# *Treponema pallidum* subsp. *pallidum* genetic population structure and relevance to syphilis prevention and treatment

**DOI:** 10.64898/2026.04.27.26351061

**Authors:** Farhang Aghakhanian, Nicole A. P. Lieberman, Christopher M. Hennelly, Jane S. Chen, Everton B. Bettin, B. Ethan Nunley, Wentao Chen, Ligang Yang, Mitch Matoga, Irving F. Hoffman, Jaime M. Altcheh, Luciana Noemí Garcia, Andrés Rabinovich, Patricia Fernández Pardal, Viviana Leiro, Ariyaratne K.A. Manathunge, Jayanthi P. Elwitigala, Sheila A Lukehart, Edward W Hook, Mauro Romero Leal Passos, Wilma Nancy Campos Arze, Hugo Boechat Andrade, Fiona Mulcahy, Qian-Qiu Wang, Rui-Li Zhang, Cai-Xia Kou, Silver K. Vargas, Kelika A. Konda, Michael Reyes Diaz, Tran Veit Ha, Le Huu Doanh, Shu-Ichi Nakayama, Yuki Ohama, Makoto Ohnishi, Bin Yang, David Šmajs, Petra Pospíšilová, Melissa J. Caimano, Jonathan J. Juliano, M. Anthony Moody, Juan C. Salazar, Justin D. Radolf, Patricia Nadal-Barón, Linda H. Xu, Lorenzo Giacani, Kelly L. Hawley, Alexander L. Greninger, Arlene C. Seña, Jonathan B. Parr

**Author notes:** Co-senior authors (Alexander L. Greninger, Arlene C. Seña, Jonathan B. Parr). Corresponding authors: Jonathan B. Parr.

## Abstract

Syphilis, caused by the spirochete *Treponema pallidum* subsp. *pallidum* (TPA), is resurging globally, particularly in low- and middle-income countries. However, TPA genomic diversity and population structure in these settings remain poorly characterized.

We investigated the global genetic diversity of syphilis spirochetes, sequencing 298 new TPA genomes from 11 countries across five continents, including underrepresented areas such as Argentina, Colombia, Malawi, Sri Lanka, and Vietnam. Combined with 1,409 public genomes, our dataset comprised 1,707 genomes. Hierarchical clustering identified six Nichols and five SS14-lineage subpopulations, with distinct subpopulations concentrated in Africa, East Asia, and the Americas, as well as previously unrecognized diversification within the globally dominant SS14 lineage. Concordance analysis showed that widely used multilocus sequencing typing methods recapitulate major Nichols-lineage subpopulations but have reduced discriminatory power for the SS14 lineage. Genome-wide Fixation index scans and targeted analyses of genes encoding outer membrane proteins prioritized for vaccine development demonstrated lineage- and subpopulation-specific patterns of genetic structure and selection. We observed strong diversifying selection acting on cell envelope assembly factors (BamA, LptD), selected FadL-like transporters, members of the *T. pallidum* repeat (Tpr) family, and efflux-associated outer membrane factors, alongside strictly conserved β-barrel scaffolds. Macrolide resistance and reduced beta-lactam susceptibility marker prevalence varied by lineage and geographical region.

These findings refine our understanding of TPA’s genetic diversity, delineate heterogeneous evolutionary trajectories across key vaccine-relevant loci, and underscore the importance of geographically representative genomic analyses to inform syphilis vaccine design and for antimicrobial resistance monitoring.

**Significance Statement:** Despite increasing syphilis rates worldwide, genomic data from its causative agent *Treponema pallidum* subsp. *pallidum* have largely originated from a small number of high-income countries. This study significantly expands our understanding of TPA genomic diversity by sampling across underrepresented low- and middle-income countries (LMICs) in Africa, Asia and South America. We analyze 1,707 genomes, including 298 newly sequenced from 11 LIMCs, and identify novel subpopulations, lineage-specific variation in outer membrane proteins, and geographic differences in antimicrobial resistance markers. These results illustrate how sampling from understudied regions reveals previously unappreciated, geographically structured diversity, improving our understanding of TPA population structure and informing the development of globally relevant vaccines, treatment strategies, and diagnostic tools.

## Introduction

Syphilis, a sexually transmitted infection caused by *Treponema pallidum* subsp. *Pallidum* (TPA), continues to present a major global health challenge. Since the turn of the twenty-first century, syphilis cases have increased globally, particularly in many low- and middle-income countries (LMICs) and, more recently, in some high-income countries (1–5). The World Health Organization (WHO) estimated 8 million cases of syphilis in 2022, with most reported in the African and American regions (6). The surge in cases and congenital syphilis in particular highlight the pressing need for improved diagnostic, preventive and therapeutic strategies to curtail the global epidemic (7).

Although continuous TPA cultivation is now possible using specialized co-culture with rabbit epithelial cells (8), isolating the organism directly from clinical specimens remains challenging. As a result, rabbit intratesticular inoculation is still widely used for laboratory propagation (9, 10). Hybrid-capture enrichment methods for TPA sequencing (11, 12) have, therefore, been pivotal, enabling whole-genome recovery directly from clinical samples and accelerating discoveries related to pathogen diversity, vaccine targets, and control strategies.

Whole-genome sequencing has revealed two distinct TPA lineages, Nichols and SS14, that circulate globally (9, 12–14), each comprising multiple subpopulations. No differences in clinical manifestations between the two lineages have been documented. However, research into the spatiotemporal dynamics and genetic variation of these subpopulations has been limited to samples from fewer than 14% of all countries. The functional importance of observed genetic variation remains largely unknown, underscoring the need for focused research to better understand how these variations contribute to disease pathogenesis and to guide future prevention and treatment approaches.

Most TPA sequences originate from high-income countries (15–19), leaving LMICs underrepresented. This disparity limits insight into TPA global genetic diversity, particularly for outer membrane proteins (OMPs) that play central roles in pathogen-host interactions and immune evasion and are considered primary vaccine targets (20, 21). Considerable OMP heterogeneity, including within the TPA repeat (Tpr) family, is well documented and pertinent to vaccine design, as well as transmission and molecular epidemiology studies (20, 22–26). Macrolide resistance markers show considerable geographical variation (14), but little is known about the distribution of penicillin-binding protein variants recently shown to reduce *in vitro* susceptibility to penicillin, the first-line treatment for syphilis, and ceftriaxone (27).

To date, most analyses of TPA genetic diversity have focused on using core genome variants to define phylogenetic relationships, thus excluding genes that undergo recombination or otherwise evolve rapidly. However, this approach limits our understanding of evolution of immune-selected outer membrane proteins, which is necessary for the development of a successful vaccine. Here, we address gaps in our understanding of TPA genetic diversity through expanded sampling and TPA genomic sequencing in partnership with research sites across three continents and employ both short- and long-read sequencing to analyze core genome and a subset of accessory genome variants. Building on prior work (13, 14, 28), we generate new genomes from undersampled regions of the world and integrate them with publicly available data to examine contemporary TPA genomic epidemiology, emphasizing population structure, genes encoding vaccine-relevant proteins, and markers of antimicrobial resistance.

## Results

### Study population

The number of complete genomes from clinical samples increased rapidly starting in 2016. However, geographic sampling remains uneven, with particularly sparse representation from West, Central, and South Asia. Coverage is even more limited across Africa and Southeast Asia, where roughly 60 countries are not represented (**Figure 1**). We generated 298 new, high-quality TPA whole-genomes from participants with primary, secondary, and early latent stages of syphilis sampled in 11 countries across five continents. Of these, 204 were obtained from participants enrolled in a cross-sectional study conducted in Argentina, China, Colombia, Malawi, Sri Lanka, and Vietnam. The remaining 94 came from existing samples previously collected during cross-sectional studies performed in Argentina, Brazil, China, Ireland, Japan, Madagascar, and Peru (**Supplementary Table 1**). We combined these new genomes with 1,409 publicly available genomes, resulting in a final analysis dataset of 1,707 genomes (**Supplementary Table 2**).

**Figure 1:**
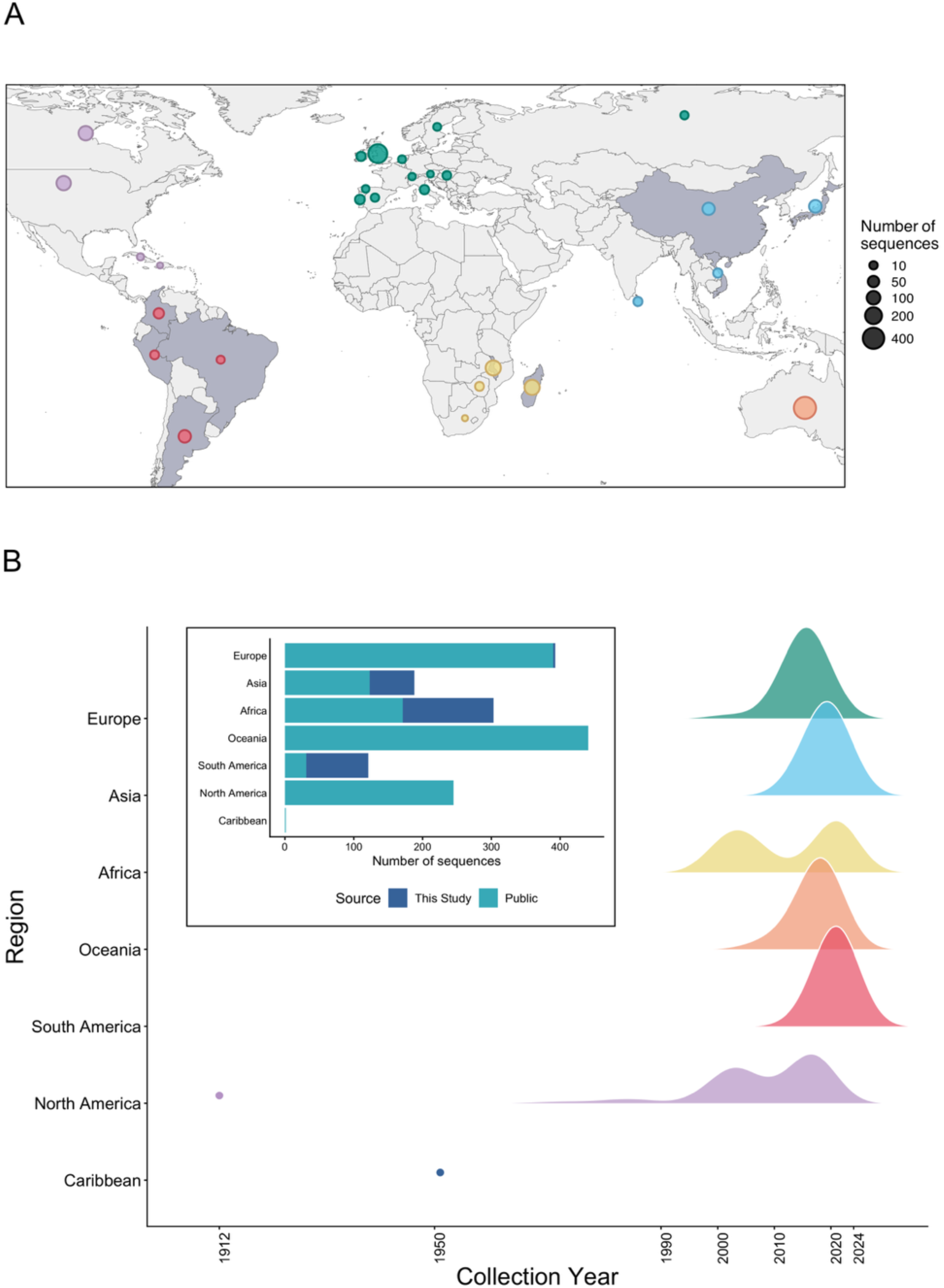
Geographic and temporal distribution of *TPA* genomes included in this study. (A) Distribution by country, with countries represented in this study shaded in gray. (B) Distribution by year of isolation and the proportion of genomes sequenced in this study relative to publicly available data across regions. The distribution highlights sparse historical sampling of TPA genomes worldwide, particularly in Asia and Africa.

### Genetic population structure by country

Of the newly sequenced isolates, 42.3% (n=126) belonged to the Nichols lineage and 57.7% (n=172) to the SS14 lineage (**Figure 2 and Supplementary Figure 1**), which differs from distribution in public data (25.3% Nichols and 74.7% SS14). In Argentina, the lineages were nearly evenly represented, with 47.1% (n = 33) Nichols and 52.9% (n = 37) SS14. All isolates from Sri Lanka (n=16), as well as the majority from Brazil (75%, n=6) and Vietnam (63.2%, n=12), belonged to the SS14 lineage. All isolates from Madagascar (n=60) were of the Nichols lineage, consistent with previous reports (28).

**Figure 2:**
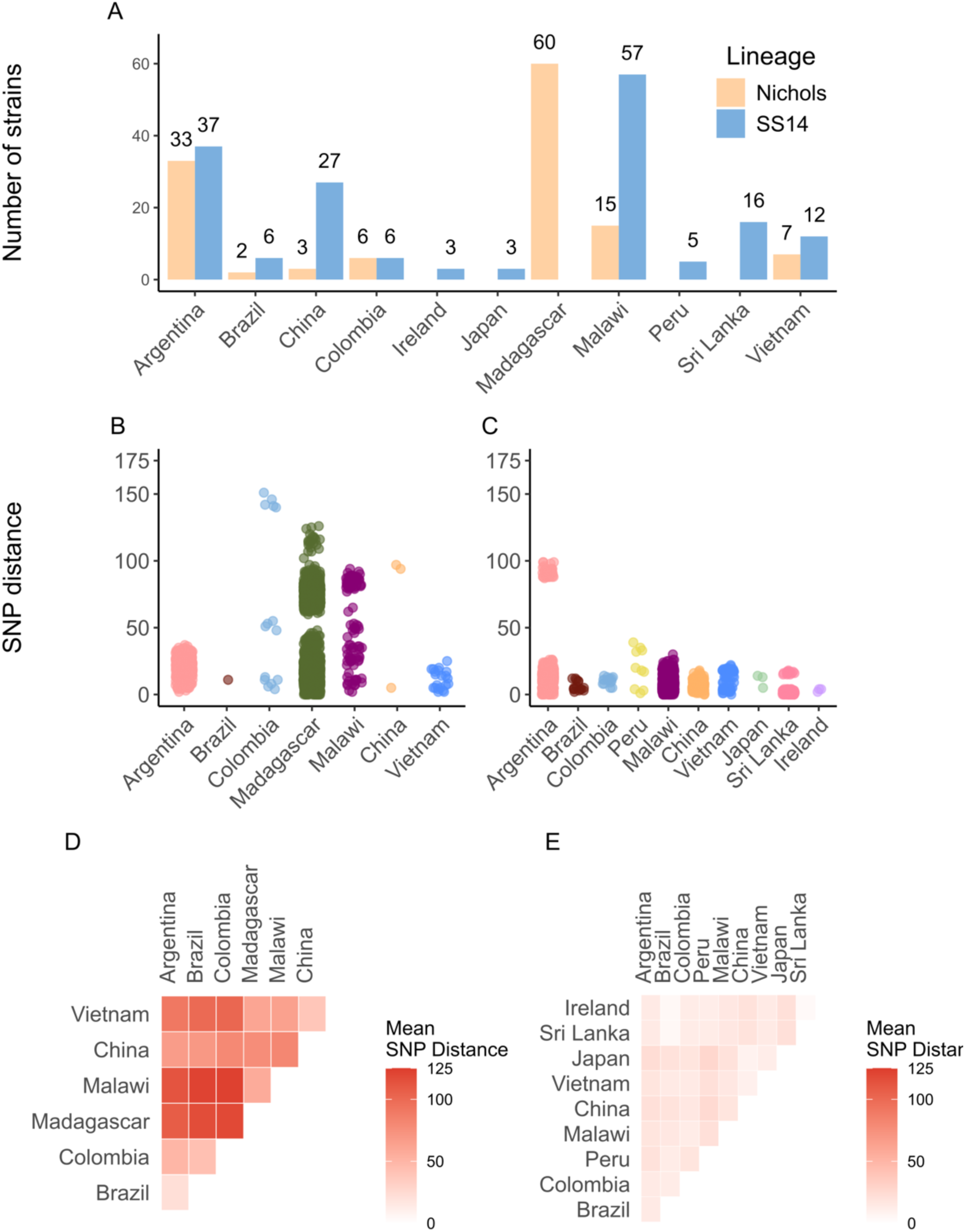
Characteristics of newly sequenced strains in this study, including the (A) number sequenced by lineage and country; within-country pairwise SNP distance distributions in the (B) Nichols and (C) SS14 lineages; and between-country average pairwise SNP distance in the (D) Nichols and (E) SS14 lineages. In Nichols-lineage genomes from Colombia, the pattern of observed pairwise SNP distances is driven in part by one genome (TPVCC052NS).

We calculated pairwise within- and between-country single nucleotide polymorphism (SNP) distances (**Figure 2B-F**) using whole-genome sequences with manual masking of the *tpr* gene family, *arp*, *tp0470*, and 23S rRNA loci. Within any one country, we found fewer than 151 and 39 pairwise SNPs differences for the Nichols and SS14 lineages, respectively, with two notable exceptions. First, among SS14-lineage strains from Argentina, we observed one sample (GSPAR082YP) that showed between 87 and 99 SNP differences (median of 91) compared to other Argentinian strains. Phylogenetic analysis revealed that this genome clusters with Mexico-A, a divergent SS14-lineage strain collected from Mexico in 1953 (**Supplementary Figure 2**) (28). In a previous study, we identified five samples from Baltimore, MD, USA, that also are closely related to Mexico-A, two of which (MD06B and MD18B) were previously sequenced (28, 29) (**Supplementary Figure 3**). Similarly, within the Nichols-lineage in Colombia, we identified an isolate, TPVCC052NS, that appears as an outgroup to the South American Nichols-lineage strains, genetically closer to Australian Nichols-lineage strains reported by Taouk *et al.* (17) (**Supplementary Figure 2**). These patterns suggest the presence of rare divergent isolates or possible introduction via international travel.

For countries with more than three strains, we calculated between-country pairwise SNP distances. Median SNP differences were 81 (interquartile range [IQR] 34-109) and 15 (IQR 11-18) for Nichols and SS14 lineages, respectively. For the Nichols lineage, the highest between country pairwise SNP distances were observed between African strains from Malawi and Madagascar and strains from South America, including those from Argentina, Brazil, and Colombia. No comparable geographic differences were observed for the SS14-lineage isolates.

### Identification of distinct TPA subpopulations

We identified six Nichols and five SS14-lineage subpopulations using Bayesian clustering analysis of manually masked data (**Figure 3**), an increase of one Nichols and two SS14 subpopulations compared with earlier analyses (14). The major circulating Nichols-lineage subpopulations were Nichols-1 and Nichols-3, which include samples from Oceania, South America and Europe. Strains from Africa were found in two subpopulations: Nichols-2 (aligns with the Nichols A subclade in Lieberman *et. al.* (28) ), including strains from Madagascar (n=92), Malawi (n=16), Zimbabwe (n=1), South Africa (n=1), and Cuba (n=1); and Nichols-4, corresponding to the Nichols B subclade in Lieberman *et. al.* (2021) and comprising genomes from Madagascar (n=51, including 20 newly sequenced for this study), Malawi (n=11), UK (n=1), Zimbabwe (n=4). Sampling year did not correlate with subpopulation assignment for strains from Madagascar, where sampling commenced in 2000 (Mann-Whitney U Test p-value = 0.73). Nichols-5 contained isolates closely related to the Nichols reference genome (CP004010.2), while Nichols-6 consisted of four isolates genetically closer to the Sea 81-4 strain.

**Figure 3:**
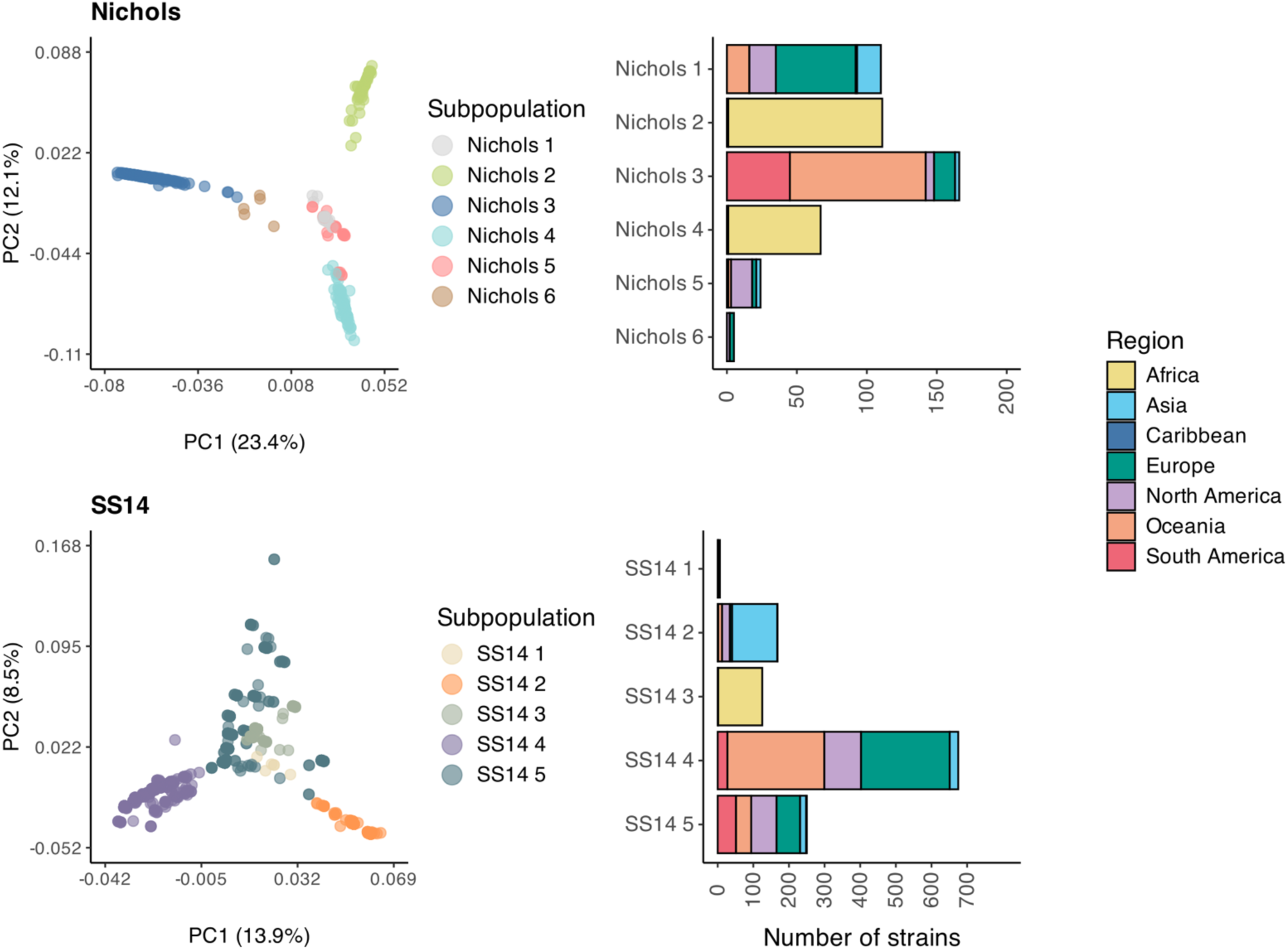
Principal component analysis (PCA) demonstrates genetic population structure within the two TPA lineages (Nichols and SS14). Analysis of sample origins reveals distinct geographical composition for some subpopulations; for example, Nichols-2, -4, and SS14-3 are composed almost entirely of isolates from Africa. PCAs for Nichols- and SS14-lineage genomes are annotated by subpopulation assignment determined using Bayesian hierarchical clustering.

The SS14-1 subpopulation consisted of Mexico-A–like isolates, while the previously defined SS14-omega subclade (28) split into four subpopulations (SS14-2 to SS14-5). SS14-2 and SS14-3 included strains largely from East Asia and Africa, respectively, whereas SS14-4 and SS14-5 included globally distributed strains. SS14-4, the dominant subpopulation, was identified across all continents except Africa. Together, these results illustrate how sampling from understudied regions reveals previously unappreciated geographically structured diversity.

Principal component analysis (PCA) and minimum spanning trees (MSTs) showed inter-lineage relationships between subpopulations, with evidence of geographical clustering (**Figure 3, Supplementary Figures 4 and 5**). Within the Nichols lineage, the first principal component (PC1) differentiated Nichols-3, which is composed primarily of genomes from South America and Oceania. PC2 distinguished two African subpopulations, Nichols-2 and Nichols-4. SS14 showed a weaker population structure. Nevertheless, PC1 partially separated strains from Asia and, to a lesser extent, Africa from other populations. This pattern was more clearly resolved in the MST analysis. Within Africa, PCA identified four clusters nested within the two dominant Nichols subpopulations. One cluster within each subpopulation was almost entirely composed of Madagascar genomes, while the other cluster contained representatives from Madagascar, Malawi, and Zimbabwe, suggesting geographically mixed transmission patterns within these secondary clusters. (**Supplementary Figure 6**).

Fixation index (F_st_) analyses identified SNPs driving subpopulation differentiation (**Supplementary Figure 7**). In the Nichols lineage, comparison between African subpopulations (Nichols-2 and 4) and South American/Oceanian subpopulation (Nichols-3), which are maximally separated in the Nichols-lineage PC1, highlighted non-synonymous, highly differentiated SNPs (F_st_ > 0.9) in *tp0462* (n=17 SNPs), a putative lipoprotein (30), and *tp0865* (n=13 SNPs), a member of the FadL outer membrane protein (OMP) family (20). Comparisons between African Nichols-lineage subpopulations revealed high-F_st_ SNPs in *tp0136* (n=7 SNPs) and *tp0483* (n=2 SNPs), both suggested to encode fibronectin-binding proteins (31, 32). In the SS14-lineage, variance along PC1 was mainly driven by non-synonymous SNPs in *tp0705 (mrcA)*. Minimum spanning trees (MST) of these genes showed distinct clustering patterns consistent with subpopulation structure, including elevated diversity in Nichols-4 for *tp0462* and *tp0865*, Africa-specific branches for *tp0136* and *tp0483*, and a distinct SS14-4 cluster for *tp0705*.

### MLST and whole-genome FastBAPS analysis concordance is strong for Nichols-but not SS14-lineage samples

A three-locus MLST scheme, based on *tp0136*, *tp0548* and *tp0705*, historically aided population structure inference prior to the availability of WGS data and remains in use in many regions (33). To evaluate the concordance of this MLST scheme with WGS, we compared MLST sequence types with WGS-based FastBAPS hierarchical clustering. The majority of African isolates carried novel alleles at one or more MLST loci that were not represented in current databases, despite clear WGS-based clustering assignment (**Supplementary Figure 8**). Therefore, concordance analyses were limited to isolates with assignable MLST types.

Within the Nichols lineage, MLST and FastBAPS showed strong agreement (adjusted Rand index [ARI] = 0.965). The dominant sequence types, ST26 (n = 153) and ST6 (n = 101), mapped almost exclusively to single FastBAPS clusters (Nichols-3 and Nichols-1), indicating that MLST effectively captures major population structure in this lineage. In contrast, the SS14 lineage showed lower concordance (ARI = 0.655). Although common sequence types ST1 (n = 493), ST3 (n = 128), and ST114 (n = 89) corresponded broadly to FastBAPS-defined subpopulations, each sequence type was distributed across multiple subpopulations, revealing finer-scale structure detectable only by WGS. For example, ST1 spanned SS14 subpopulations 1–4, and ST3 spanned subpopulations 2 and 5. This reduced discriminatory power (reflected in lower ARI and greater cross-partitioning) likely stems from both lower nucleotide diversity within the SS14 lineage and inherent limitations of a three-locus scheme. Together, these results demonstrate that while MLST provides a reliable framework for Nichols lineage classification, WGS-based FastBAPS clustering offers superior resolution in genetically diverse populations and captures population structure undetected by the conventional three-locus MLST system.

### Genetic diversity and evolutionary signatures in non-Tpr outer membrane proteins

In this study, the non-Tpr OMPs comprise two stand-alone OMP assembly machinery (BamA and LptD), long-chain-fatty-acid transporters (FadLs), and 8-stranded β-barrels and OM factors (OMFs) for efflux pumps (20). Our analysis reveals a heterogeneous evolutionary landscape shaped by disparate selective pressures (**Supplementary Table 3**). OMP profiles were strongly associated with SS14 vs. Nichols lineage membership (**Supplementary Figure 9**). The core OMP assembly machinery *tp0326* (*bamA*) and *tp0515* (*lptD*) showed clear evidence of diversifying selection by the Branch-Site Unrestricted Statistical Test for Episodic Diversification (BUSTED) (p = 2.46×10⁻⁹ and 3.00×10⁻⁴, respectively), along with elevated haplotype and nucleotide diversity, indicating that even essential, functionally constrained proteins undergo diversification likely driven by host immune pressure.

The FadL-like family displayed the most pronounced heterogeneity, and *tp0548* emerged as the most diverse gene (haplotype diversity (Hd) = 0.820; nucleotide diversity (π) = 0.00996). Given that TPA is a fatty acid auxotroph that relies heavily on host-derived lipids (30), the extreme diversification of *tp0548* suggests it may function as a high-affinity scavenger operating at the host-pathogen interface, where it is driven to alter its extracellular loops to evade neutralizing antibodies. Three additional FadL homologs, *tp0856*, *tp0859*, and *tp0865*, also exhibited diversifying selection signals. *tp0865* showed high nucleotide diversity similar to *tp0548*, whereas *tp0856* and *tp0859* exhibited low nucleotide diversity (π = 6.01×10^-5^ and π = 3.32×10^-5^, respectively) despite diversifying selection signals. Our earlier studies showed that the barrel structure of FadLs is highly constrained, but mutations accumulate in their immunogenic extracellular loops (ECLs) (26, 34). Consistent with our previous results, the pattern in *tp0856* and *tp0859* suggests that only a few ECLs are under positive selection. *tp0858* accumulated many segregating sites (n = 195) and had a large theta (θ = 24.3), but lacked evidence of diversifying selection, implying either balancing selection or near-neutral evolution at this locus. These patterns indicate that FadL-like genes follow distinct evolutionary strategies, with some (*tp0548* and *tp0865*) showing evidence of positive selection in multiple extracellular loops and others *(tp0856* and *tp0859*) exhibiting positive selection restricted to only a few extracellular loops.

All genes encoding 8-stranded β-barrels (*tp0126*, *tp0479, tp0698,* and *tp0733*) exhibit very low genetic diversity, with minimal nucleotide diversity and no diversifying selection signals, consistent with essential structural roles. Genes encoding outer membrane factors for efflux pumps (*tp0966*, *tp0967*, *tp0968*, and *tp0969*) showed heterogeneous evolutionary patterns. *tp0966* displayed moderate haplotype diversity and significant diversifying selection (p = 1.00×10^-4^). This pattern suggests that *tp0966* may interact with the host environment or transport diverse substrates, subjecting it to positive selection. *tp0968* also showed diversifying selection but with lower background diversity (Hd = 0.054; π = 4.68×10^-5^), paralleling patterns of localized epitope variation under structural constraint. In contrast, *tp0967* exhibited moderate haplotype diversity (Hd = 0.497) but low nucleotide diversity (π = 1.22×10^-4^) and no selection signal, suggesting evolution under purifying selection. *tp0969* showed an unusual profile, with high haplotype diversity (Hd = 0.645) but low nucleotide diversity (π = 9.69×10^-5^) and 29 segregating sites. This pattern may reflect recent population expansion or retention of older haplotypes rather than ongoing adaptation. Together, the efflux system genes do not appear to be under uniform selective pressure, suggesting functional specialization or differential importance in TPA pathogenesis.

### Genetic diversity in Tpr gene families

The genetic diversity of TPA is heavily driven by the hypervariable *tpr* family, which is challenging to resolve using short-read WGS data. Therefore, targeted Oxford Nanopore Technologies (ONT) sequencing was conducted on seven TPA *tpr* genes, encompassing both subfamily I (*tp0117*, *tp0131*, *tp0316*, *tp0620*) and subfamily II (*tp0313*, *tp0317*, *tp0621*). *tprF* (*tp0316*) was excluded from analysis due to gene truncation in TPA (35). Among the remaining *tpr* genes, calling success varied by gene, ranging from 97.6% for *tprD* (*tp0131*) to 43.2% for *tprI* (*tp0620*). Failure to resolve *tp0620* was attributed to its complex tandem repeat structure and homopolymer-rich regions that pose challenges for ONT basecalling. Of 206 samples on which targeted *tpr* sequencing was attempted, 84 samples (40.8%) yielded successful assemblies for all seven *tpr* genes.

Among the six *tpr* genes analyzed, subfamily II exhibited substantially higher diversity than subfamily I (**Figure 4, Supplementary Figures 10 and 11**, and **Supplementary Table 4**). BUSTED analysis confirmed that all six *tpr* genes show signatures of episodic diversifying selection, indicating that immune pressure and/or functional divergence have driven adaptive evolution across both subfamilies. Within subfamily I, *tprD* (*tp0131*) was the most diverse gene (π = 0.011, 247 segregating sites, θ = 42.02), with an accumulation of mutations throughout its central variable region (CVR) and discrete sites in C-terminal. Within subfamily II, *tprJ* (*tp0621*) stood out as the most hypervariable locus in the analyzed *tpr* repertoire (π = 0.051, 441 segregating sites, θ = 75.35, Hd = 0.78).

**Figure 4:**
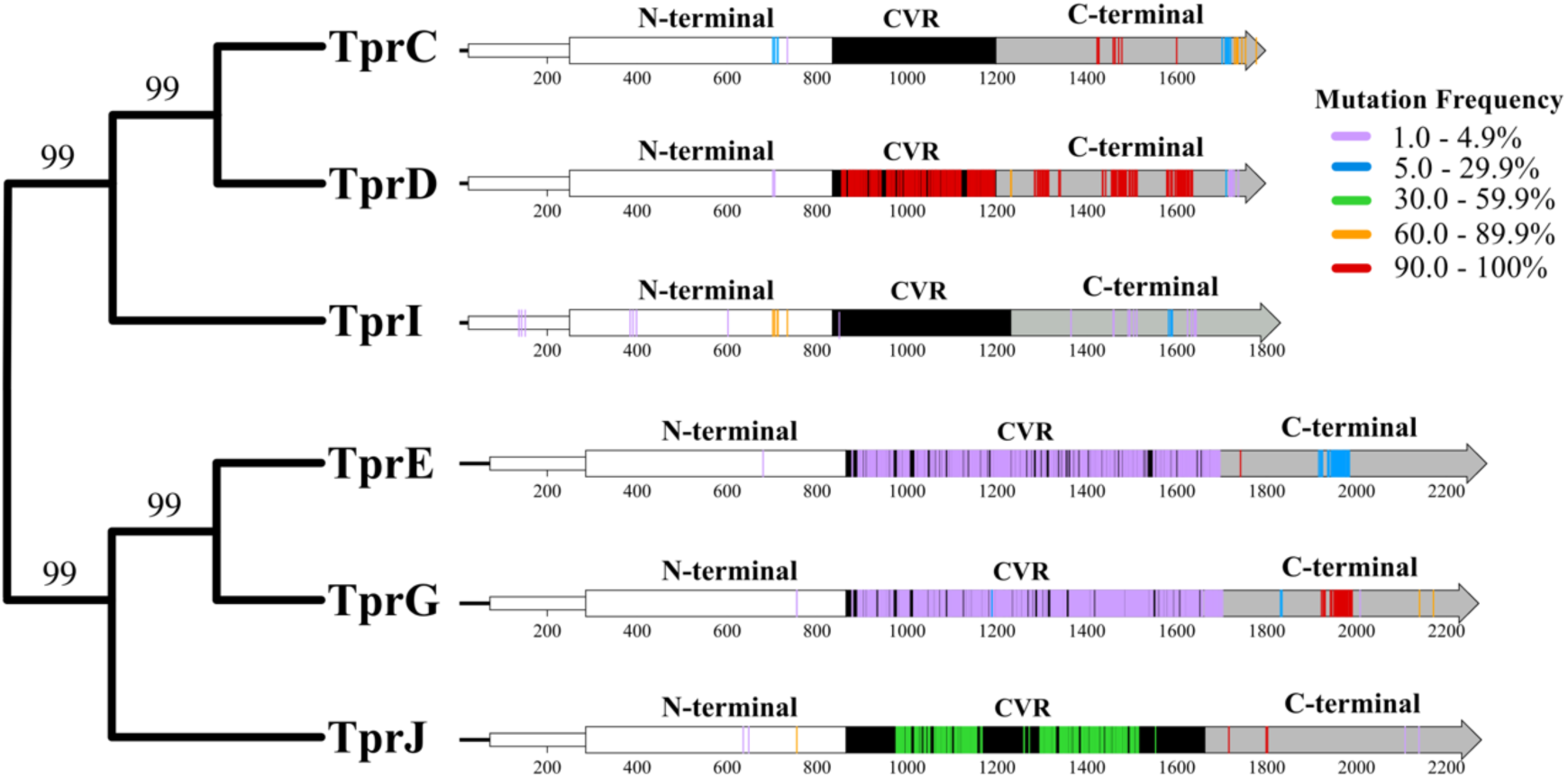
Phylogenetic reconstruction of *tpr* subfamilies I and II using consensus sequences from the Nichols (CP004010.2) reference genome. The right panel displays results from targeted ONT sequencing of 206 samples from China, Vietnam, Colombia, and Malawi as part of this study. SNPs are mapped to the linear Tpr structures and color-coded according to allele frequency (AF) compared to reference.

Allele frequency spectra revealed distinct evolutionary dynamics between structural domains and subfamilies. Overall variation in the N-terminal region of all Tpr proteins was minimal. Within subfamily I, the CVR regions of *tp0117* (*tprC*) and *tp0620* (*tprI*) were highly conserved, with mutations present at very low frequency and restricted to a few discrete sites. In contrast, *tp0131* (*tprD*) showed a dramatically different pattern, with high-frequency variants compared to the reference (30–100% allele frequency) densely distributed throughout the CVR. Earlier studies reported two main *tprD* alleles in TPA: a *tprD* allele found in the Nichols and Chicago strains and a distinct *tprD2* allele found in the SS14, MexicoA, Sea81-4, and Bal3 strains (25, 36). In contrast, our analysis showed that the majority of contemporary sequences, regardless of lineage, had alleles genetically more similar to *tprD2* than to the Nichols *tprD* allele. The only exception was the Nichols-4 subpopulation, in which all isolates carried a *tprD* allele that is more closely related to the Nichols reference.

For subfamily II, the CVRs were predominantly characterized by low-frequency variants (1–5% allele frequency) in both *tp0313* (*tprE*) and *tp0317* (*tprG*). This contrasts sharply with *tp0621* (*tprJ*), where the CVR exhibited intermediate- to high-frequency variants (30–60% allele frequency), indicating that *tprJ* polymorphisms have risen to appreciable frequency and were likely maintained by balancing selection, immune-driven frequency-dependent selection, or geographic/temporal population structure. In the C-terminal, mutations were similarly restricted to discrete regions across most *tpr* genes.

Elevated *tprJ* allele frequencies may reflect its role encoding a B-cell target (37) with multiple co-circulating antigenic variants, analogous to the antigenic variation system of TprK (38) but operating through standing genetic variation rather than localized gene conversion from donor cassettes. Alternatively, the high-frequency TprJ variants could represent functional alleles adapted to different host immune backgrounds or tissue microenvironments, particularly if TprJ surface regions mediate immune evasion, epitope masking, or nutrient acquisition in addition to serving as targets for B-cell recognition. The combination of high nucleotide diversity, numerous segregating sites, elevated haplotype diversity, and intermediate-frequency alleles in the CVR uniquely positions *tprJ* as a dynamically evolving *tpr* locus (excluding *tprK*, which was not analyzed here).

### Antibiotic resistance markers

We identified three highly differentiated non-synonymous SNPs in penicillin-binding protein TP0705 (MrcA): C1517T, A1873G, and G2122A corresponding to A506V, M625V, and G708S, respectively. Recent in vitro work by Pospíšilová *et al*. (27) found that M625V reduces susceptibility to ceftriaxone and penicillin G, although no clinical treatment failures have been reported (39). A1873G (M625V) occurred exclusively in SS14-lineage isolates and was particularly prevalent in SS14-4 and SS14-5 subpopulations; 629 SS14-4 isolates (93.2%, 95% CI: 91.0% – 94.9%) and 160 SS14 5 (64%, 95% CI: 57.9% – 69.7%) carried this variant. Nearly all isolates with A1873G also harbored G2122A (n=781, 99.0% 95% CI: 98.0% – 99.5%). The C1517T was frequent (n=138, 82.7% 95% CI: 76.2% - 88.1 %) in SS14-2 (**Supplementary Figure 12**).

Macrolide resistance marker (23S rRNA A2058G or A2059G) prevalence varied starkly by geography, ranging from 100% in all sampled Asian countries (China, Japan, Vietnam, and Sri Lanka) and in Peru to very low in Africa, with only 1.6% prevalence in Madagascar (95% CI: 0.04%–8.94%) and 6.9% in Malawi (95% CI: 2.3%–15.5%). Lineage-specific analysis revealed a significant difference only in Argentina, where macrolide resistance was markedly higher in SS14-lineage strains (51.4%, 95% CI: 34.4%–68.1%) compared with Nichols-lineage strains (15.2%, 95% CI: 5.1%–31.9%; Fisher’s exact test, P = 0.002).

## Discussion

Our analysis of nearly 300 new whole-genome sequences from historically undersampled LMICs expands understanding of contemporary TPA strains circulating worldwide. In line with previous work (12–14), we confirm greater genetic diversity in the Nichols than the SS14 lineage. We identify eleven global subpopulations, including three composed almost exclusively of genomes from Africa, and likely a reflection of expanded sampling. Targeted long-read sequencing of the *tpr* gene family complemented genomic analysis and enabled deeper exploration of genetic diversity across TPA outer membrane proteins (OMPs). We identify clear subpopulations and geographical differences in OMP diversity relevant to the development of a globally effective syphilis vaccine, as well as regional variation in the prevalence of AMR-associated mutations. These findings provide new insights into TPA genetic epidemiology with direct relevance to syphilis prevention and treatment efforts.

Our expanded dataset refines the global population structure of TPA and highlights specific genes that may underlie geographic clustering and merit further functional study. For example, we identify two Nichols-like subpopulations in Africa: Nichols-4, composed exclusively of genomes from Madagascar, and Nichols-2, which includes genomes from Madagascar, Malawi, South Africa, and Zimbabwe. Analyses of Madagascar samples collected over time do not support collection date as a major driver of subpopulation assignment. Variation in fibronectin-binding proteins TP0136 and TP0483, which have been implicated in attachment to host cells (31, 32, 40, 41) and show evidence of recombination and positive selection (40, 42), appears to be a key feature distinguishing Nichols-2 and Nichols-4 subpopulations.

Within the Nichols lineage, the Nichols-3 subpopulation (predominantly from South America and Oceania) shows clear segregation from other worldwide strains, largely driven by strikingly elevated diversity in *tp0462* and *tp0865*, both of which are subject to recombination and selection and may experience host immune pressure (42, 43). Nichols-3 harbors 78 distinct *tp0462* haplotypes (74 proteoforms) compared to only 9 haplotypes (8 proteoforms) in all other subpopulations combined, and 26 distinct *tp0865* haplotypes (25 proteoforms) versus 10 haplotypes (10 proteoforms) elsewhere, although the functional drivers and clinical implications of this localized diversification remain unclear.

MLST has been widely used for epidemiological surveillance of TPA. Our analysis shows that the established three-locus MLST scheme’s performance varies substantially by lineage. Within the Nichols lineage, MLST results align well with population structure inferred by FastBAPS analysis of WGS data, indicating that the current scheme sufficiently resolves broad subdivisions in this group. In contrast, MLST performs poorly for the globally dominant SS14 lineage, likely due to its lower nucleotide diversity and the limited discriminatory capacity of the three-locus scheme for recently diverged strains. Inclusion of additional polymorphic loci, particularly those exhibiting elevated nucleotide diversity or evidence of diversifying selection within SS14, would improve resolution for the SS14 lineage and strengthen the epidemiological utility of MLST.

Our analysis of the non-Tpr OMPs indicates that TPA evolutionary dynamics extend beyond the evolutionary plastic *tpr* family, with other OMPs also being actively shaped by host immune pressure. BamA (TP0326) and the LptD homologue TP0515 both show strong signatures of diversifying selection, consistent with prior evidence that surface-exposed loops of BamA and other OMPs are targets of opsonic antibodies (44–46). BamA, which is essential for OMP biogenesis, concentrates its variation in extracellular loops that segregate by geography and TPA subpopulation (26), suggesting lineage-specific adaptation to local immune landscapes. Likewise, the retention and diversification of TP0515 in a spirochete lacking classical lipopolysaccharide implies a repurposed but immunologically exposed role, potentially in maintaining outer membrane asymmetry or transporting alternative glycolipids (20).

Within the FadL-like family, evolutionary signatures are highly heterogeneous. TP0548 displays extreme diversity and lineage-specific segregation, reflecting a presumably metabolically essential yet immunologically exposed protein under strong local selection. In contrast, subgroup II paralogs TP0856/TP0858 and subgroup III paralogs TP0859/TP0865 displayed differential diversification, where one member of each subgroup is relatively conserved while the other paralog has diversified. The conserved proteins might maintain core transport roles under episodic selection pressure, while their more variable counterparts could buffer adaptive evolutionary changes. Interestingly, the highly variable locus TP0858 encodes a more abundant TP0858 protein in the TPA proteome than TP0856 (47) and has been identified as a gene-conversion acceptor locus (48). Comprehensive structural, functional, and immunological studies of individual family members will be required to elucidate the mechanisms underlying these distinct evolutionary patterns. All 8-stranded β-barrels remain nearly invariant and poorly immunogenic (34), while selected efflux-associated OMPs (especially TP0966, with strong serological recognition and diversifying selection) bear site-specific signatures of selection. Together, these patterns indicate that only a subset of the TPA surface is heavily engaged in immune and ecological interactions, whereas the remainder is constrained to preserve core outer membrane architecture and essential transport.

Tpr analyses suggest that different members of the repertoire occupy distinct evolutionary “niches,” with some genes (for example, *tprC*, *tprI*) remaining relatively conserved while others (particularly *tprJ*) accumulate extensive standing variation under episodic diversifying selection. The apparent predominance of *tprD2-*like alleles, with Nichols-like *tprD* largely confined to Nichols-4, further points to a selective regime that shapes which variants become fixed or persist at high frequency. It is important to note that the *tprK* gene plays an essential role in *T. pallidum* immune evasion but was not included in this analysis due to challenges of assembling its hypervariable regions using the sequencing approaches employed here. Higher fidelity long-read sequencing approaches have recently been deployed to enable in-depth investigation of *tprK* diversity and spirochete transmission (49).

While this study includes the largest TPA genomic dataset analyzed to-date, there are several limitations. First, the generalizability of our findings remains limited by sparse TPA genomic data across many regions of the world. Additional sequencing in countries not represented in TPA genomic databases is particularly important for the design of a globally effective syphilis vaccine. Discovery of new TPA subpopulations in Africa with distinct OMP variants underscores the value of expanding syphilis research to include undersampled regions. Second, we experienced technical challenges with high-quality *tprK* amplicon ONT sequencing, which limited our ability to fully resolve the diversity of this important gene’s hypervariable regions with this platform. Third, the absence of experimentally-determined structures for the Tpr family prevented us from mapping identified sequence variation onto specific structural domains, limiting our ability to investigate the evolutionary drivers of these changes. This limitation underscores the need to determine high-confidence Tpr protein structures. Finally, genetic markers of antibiotic resistance do not always translate to meaningful phenotypes. We identify differences in the geographical distribution of macrolide resistance markers, found at high frequency worldwide with the exception of Malawi, Madagascar, and Argentina, and consistent with prior reports (14). We also identify a recently reported marker of decreased in vitro penicillin susceptibility exclusively in the globally dominant SS14 lineage; however, the significance of this finding is unclear in the absence of clinical failure, which has not been documented in the affected countries.

Together, these findings refine our understanding of contemporary TPA genomic epidemiology by revealing previously unrecognized subpopulations and linking them to distinct variation in candidate outer membrane vaccine targets and antimicrobial resistance markers. Our findings demonstrate that TPA lineages and subpopulations vary not only in their core genome but also in the diversity of surface-exposed proteins that underpin ongoing vaccine development. This heterogeneity highlights opportunities to integrate genomic analysis with immunological and functional studies to define which variants are most relevant for TPA transmission, pathogenesis, and immune escape. Continued expansion of genomic sampling, particularly in underrepresented regions, will be essential to capture the full spectrum of circulating diversity, support vaccine design, and better inform global syphilis prevention and control efforts.

## Materials and Methods

This study included both active enrollment of participants as part of a multicenter, cross-sectional study and use of banked clinical samples from individuals with early syphilis across sites in South America, Africa, Asia, and Europe, under protocols approved by institutional review boards at the University of North Carolina, the University of Washington, and participating countries. Written informed consent was obtained from all participants, and specimens were de-identified prior to analysis. Detailed ethical approvals and site-specific protocols are described in the **Supplementary Materials**.

In brief, samples were processed and analyzed as follows. Genomic DNA was extracted from lesion swabs, biopsies, blood, and rabbit-passaged isolates, enriched for TPA using hybrid capture, and sequenced on Illumina platforms at UNC and UW. Reads underwent stringent quality control, host DNA filtering, mapping to Nichols and SS14 reference genomes, and standardized variant calling; we applied the same pipeline to newly generated genomes and publicly available data. Targeted long-read sequencing with Oxford Nanopore was used to characterize selected *tpr* family loci, using nested PCR, MinION sequencing, and consensus assembly with established long-read polishing tools. Full methodological details, including primer schemes, enrichment conditions, and all software parameters are provided in the Supplementary Materials.

Population genomic analyses were based on SNPs outside highly repetitive and difficult-to-map regions and included lineage-specific clustering (FastBAPS) (50), phylogenetic inference (IQ-TREE 2) (51), and visualization in R (52). We quantified population structure and differentiation using PCA, pairwise SNP distances, and F_st_, annotated variants with SnpEff (53), and assigned sequence types using the TPA MLST scheme (33) and PubMLST (54). For OMP genes, we estimated standard diversity metrics and tested for episodic diversifying selection with BUSTED (55) in HyPhy (56); macrolide resistance mutations were identified using competitive mapping as previously described (14). Full details of all analytical steps, thresholds, and software versions are provided in the Supplementary Materials.

## Data Availability

All whole-genome sequencing data from this study, with residual human reads removed, are available in the Sequence Read Archive (SRA) under BioProject PRJNA815321.

## Acknowledgments

The authors express their gratitude to study participants, as well as research, clinical, and administrative staff across clinical sites. The authors also thank Viviana Leiro and Patricia Fernandez Pardal from Servicio de Dermatologia, Hospital Muñiz, Buenos Aires for their contributions.

The authors used an artificial intelligence language model for English language editing during the writing process. However, the manuscript is original to the authors, who take responsibility for its content.

## Data and code availability

Whole-genome sequencing data from this study, with residual human reads removed, are available in the Sequence Read Archive (SRA) under BioProject PRJNA815321. Analysis code is accessible at https://github.com/IDEELResearch/tpallidum-genomic-pop-structure.

## Author Contributions

The study was designed and conceptualized by ACS, ALG, JBP. Laboratory work and sequencing were performed by CMH and BEN. FA and NAPL were responsible for data curation and genomic analysis. All other authors contributed to field work, specimen collection, and interpretation of data. FA wrote the first draft of the manuscript, with input from NAPL, ALG, and JBP. All authors reviewed the final manuscript.

## References

1. R.W. Peeling, et al., Syphilis. Nat. Rev. Dis. Primers 3, 17073 (2017).

2. E.L. Korenromp, et al., Syphilis prevalence trends in adult women in 132 countries - estimations using the Spectrum Sexually Transmitted Infections model. Sci. Rep. 8, 11503 (2018).

3. K.G. Ghanem, S. Ram, P.A. Rice, The modern epidemic of syphilis. N. Engl. J. Med. 382, 845–854 (2020).

4. G. Spiteri, M. Unemo, O. Mårdh, A.J. Amato-Gauci, The resurgence of syphilis in high-income countries in the 2000s: a focus on Europe. Epidemiol. Infect. 147, e143 (2019).

5. A. Komori, H. Mori, W. Xie, S. Valenti, T. Naito, Rapid resurgence of syphilis in Japan after the COVID-19 pandemic: A descriptive study. PLoS ONE 19, e0298288 (2024).

6. World Health Organization, Global Sexually Transmitted Infections Programme. World Health Organization (2024) Available at https://www.who.int/teams/global-hiv-hepatitis-and-stis-programmes/stis/overview. Accessed 14 May 2024.

7. P. Moseley, et al., Resurgence of congenital syphilis: new strategies against an old foe. Lancet Infect. Dis. 24, e24–e35 (2024).

8. D.G. Edmondson, S.J. Norris, B.D. De Lay, Procedures for In Vitro Cultivation of Treponema pallidum, the Syphilis Spirochete. J. Vis. Exp. doi: 10.3791/66880 (2025) doi:10.3791/66880.

9. L. Yang, et al., Clinical presentation of early syphilis and genomic sequences of Treponema pallidum strains in patient specimens and isolates obtained by rabbit inoculation. J. Infect. Dis. doi: 10.1093/infdis/jiae322 (2024) doi:10.1093/infdis/jiae322.

10. J.C. Salazar, et al., Treponema pallidum genetic diversity and its implications for targeted vaccine development: A cross-sectional study of early syphilis cases in Southwestern Colombia. PLoS ONE 19, e0307600 (2024).

11. M. Pinto, et al., Genome-scale analysis of the non-cultivable Treponema pallidum reveals extensive within-patient genetic variation. Nat. Microbiol. 2, 16190 (2016).

12. N. Arora, et al., Origin of modern syphilis and emergence of a pandemic Treponema pallidum cluster. Nat. Microbiol. 2, 16245 (2016).

13. M.A. Beale, et al., Global phylogeny of Treponema pallidum lineages reveals recent expansion and spread of contemporary syphilis. Nat. Microbiol. 6, 1549–1560 (2021).

14. A.C. Seña, et al., Clinical and genomic diversity of Treponema pallidum subspecies pallidum to inform vaccine research: an international, molecular epidemiology study. Lancet Microbe 5, 100871 (2024).

15. S. Nishiki, K. Lee, M. Kanai, S.I. Nakayama, M. Ohnishi, Phylogenetic and genetic characterization of Treponema pallidum strains from syphilis patients in Japan by whole-genome sequence analysis from global perspectives. Sci. Rep. 11, 3154 (2021).

16. W. Chen, et al., Analysis of Treponema pallidum Strains From China Using Improved Methods for Whole-Genome Sequencing From Primary Syphilis Chancres. J. Infect. Dis. 223, 848–853 (2021).

17. M.L. Taouk, et al., Characterisation of Treponema pallidum lineages within the contemporary syphilis outbreak in Australia: a genomic epidemiological analysis. Lancet Microbe 3, e417–e426 (2022).

18. M.A. Beale, et al., Genomic epidemiology of syphilis in England: a population-based study. Lancet Microbe 4, e770–e780 (2023).

19. N.A.P. Lieberman, et al., Genomic Epidemiology of Treponema pallidum and Circulation of Strains With Diminished tprK Antigen Variation Capability in Seattle, 2021-2022. J. Infect. Dis. 229, 866–875 (2024).

20. K.L. Hawley, et al., Structural Modeling of the Treponema pallidum Outer Membrane Protein Repertoire: a Road Map for Deconvolution of Syphilis Pathogenesis and Development of a Syphilis Vaccine. J. Bacteriol. 203, e0008221 (2021).

21. J. Chen, J. Huang, Z. Liu, Y. Xie, Treponema pallidum outer membrane proteins: current status and prospects. Pathog. Dis. 80, (2022) doi:10.1093/femspd/ftac023.

22. C. Ávila-Nieto, et al., Syphilis vaccine: challenges, controversies and opportunities. Front. Immunol. 14, 1126170 (2023).

23. A. Addetia, et al., Comparative genomics and full-length Tprk profiling of Treponema pallidum subsp. pallidum reinfection. PLoS Negl. Trop. Dis. 14, e0007921 (2020).

24. A. Addetia, et al., Estimation of Full-Length TprK Diversity in Treponema pallidum subsp. pallidum. MBio 11, (2020) doi:10.1128/mBio.02726-20.

25. A. Centurion-Lara, et al., Fine analysis of genetic diversity of the tpr gene family among treponemal species, subspecies and strains. PLoS Negl. Trop. Dis. 7, e2222 (2013).

26. E.B. Bettin, et al., Sequence variability of BamA and FadL candidate vaccinogens suggests divergent evolutionary paths of Treponema pallidum outer membrane proteins. J. Bacteriol. 207, e0015925 (2025).

27. P. Pospíšilová, et al., Resistance to ceftriaxone and penicillin G among contemporary syphilis strains confirmed by natural in vitro mutagenesis. Commun Med (London) 5, 224 (2025).

28. N.A.P. Lieberman, et al., Treponema pallidum genome sequencing from six continents reveals variability in vaccine candidate genes and dominance of Nichols clade strains in Madagascar. PLoS Negl. Trop. Dis. 15, e0010063 (2021).

29. S.A. Lukehart, et al., Macrolide resistance in Treponema pallidum in the United States and Ireland. N. Engl. J. Med. 351, 154–158 (2004).

30. J.D. Radolf, et al., Treponema pallidum, the syphilis spirochete: making a living as a stealth pathogen. Nat. Rev. Microbiol. 14, 744–759 (2016).

31. M.B. Brinkman, et al., A novel Treponema pallidum antigen, TP0136, is an outer membrane protein that binds human fibronectin. Infect. Immun. 76, 1848–1857 (2008).

32. C.E. Cameron, E.L. Brown, J.M.Y. Kuroiwa, L.M. Schnapp, N.L. Brouwer, Treponema pallidum fibronectin-binding proteins. J. Bacteriol. 186, 7019–7022 (2004).

33. L. Grillová, et al., Molecular characterization of Treponema pallidum subsp. pallidum in Switzerland and France with a new multilocus sequence typing scheme. PLoS ONE 13, e0200773 (2018).

34. K.N. Delgado, et al., Immunodominant extracellular loops of Treponema pallidum FadL outer membrane proteins elicit antibodies with opsonic and growth-inhibitory activities. PLoS Pathog. 20, e1012443 (2024).

35. L. Giacani, K. Hevner, A. Centurion-Lara, Gene organization and transcriptional analysis of the tprJ, tprI, tprG, and tprF loci in Treponema pallidum strains Nichols and Sea 81-4. J. Bacteriol. 187, 6084–6093 (2005).

36. B. Molini, et al., B-Cell Epitope Mapping of TprC and TprD Variants of Treponema pallidum Subspecies Informs Vaccine Development for Human Treponematoses. Front. Immunol. 13, 862491 (2022).

37. B.T. Leader, et al., Antibody responses elicited against the Treponema pallidum repeat proteins differ during infection with different isolates of Treponema pallidum subsp. pallidum. Infect. Immun. 71, 6054–6057 (2003).

38. A. Centurion-Lara, et al., Gene conversion: a mechanism for generation of heterogeneity in the tprK gene of Treponema pallidum during infection. Mol. Microbiol. 52, 1579–1596 (2004).

39. L.C. Tantalo, K.D. Chamakuri, A.L. Greninger, N.A.P. Lieberman, L. Giacani, Susceptibility to Penicillin G and Ceftriaxone in Three Clinical Treponema pallidum Isolates is not Altered by Amino Acid Polymorphisms in the Tp0705 Penicillin Binding Protein. Sex. Transm. Dis. doi: 10.1097/OLQ.0000000000002291 (2025) doi:10.1097/OLQ.0000000000002291.

40. W. Li, et al., Treponema pallidum protein Tp0136 promotes angiogenesis to facilitate the dissemination of Treponema pallidum. Emerg. Microbes Infect. 13, 2382236 (2024).

41. W. Ke, B.J. Molini, S.A. Lukehart, L. Giacani, Treponema pallidum subsp. pallidum TP0136 protein is heterogeneous among isolates and binds cellular and plasma fibronectin via its NH2-terminal end. PLoS Negl. Trop. Dis. 9, e0003662 (2015).

42. M. Pla-Díaz, et al., Evolutionary Processes in the Emergence and Recent Spread of the Syphilis Agent, Treponema pallidum. Mol. Biol. Evol. 39, (2022) doi:10.1093/molbev/msab318.

43. N.A.P. Lieberman, et al., Disseminated Syphilis Caused by Two Recombining *Treponema pallidum* Strains. N. Engl. J. Med. 392, 1551–1553 (2025).

44. K.N. Delgado, et al., Extracellular Loops of the Treponema pallidum FadL Orthologs TP0856 and TP0858 Elicit IgG Antibodies and IgG+-Specific B-Cells in the Rabbit Model of Experimental Syphilis. MBio 13, e0163922 (2022).

45. K.N. Delgado, et al., Antibodies directed against extracellular loops of FadL orthologs disrupt outer membrane integrity and neutralize infectivity of Treponema pallidum, the syphilis spirochete. Frontiers in Immunology (2025).

46. A. Luthra, et al., A Homology Model Reveals Novel Structural Features and an Immunodominant Surface Loop/Opsonic Target in the Treponema pallidum BamA Ortholog TP_0326. J. Bacteriol. 197, 1906–1920 (2015).

47. J. Bosák, et al., Proteome Analysis of Seven Treponema pallidum subsp. pallidum Strains Grown In Vitro. J. Proteome Res. 24, 6091–6100 (2025).

48. M. Strouhal, et al., Complete genome sequences of two strains of Treponema pallidum subsp. pertenue from Indonesia: Modular structure of several treponemal genes. PLoS Negl. Trop. Dis. 12, e0006867 (2018).

49. N.A.P. Lieberman, et al., Lineage-specific tprK diversification and Treponema pallidum transmission dynamics in Buenos Aires, Argentina. BioRxiv doi: 10.64898/2026.01.29.702707 (2026) doi:10.64898/2026.01.29.702707.

50. G. Tonkin-Hill, J.A. Lees, S.D. Bentley, S.D.W. Frost, J. Corander, Fast hierarchical Bayesian analysis of population structure. Nucleic Acids Res. 47, 5539–5549 (2019).

51. B.Q. Minh, et al., IQ-TREE 2: New models and efficient methods for phylogenetic inference in the genomic era. Mol. Biol. Evol. 37, 1530–1534 (2020).

52. R Core Team, R: A Language and Environment for Statistical Computing (R Foundation for Statistical Computing, Vienna, Austria, 2023).

53. P. Cingolani, et al., A program for annotating and predicting the effects of single nucleotide polymorphisms, SnpEff: SNPs in the genome of *Drosophila melanogaster* strain w1118; iso-2; iso-3. Fly (Austin) 6, 80–92 (2012).

54. GitHub - tseemann/mlst: :id: Scan contig files against PubMLST typing schemes, Available at https://github.com/tseemann/mlst. Accessed 23 June 2025.

55. B. Murrell, et al., Gene-wide identification of episodic selection. Mol. Biol. Evol. 32, 1365–1371 (2015).

56. S.L.K. Pond, S.D.W. Frost, S.V. Muse, HyPhy: hypothesis testing using phylogenies. Bioinformatics 21, 676–679 (2005).

