## Supplementary Methods and Figures for "*Treponema pallidum* subsp. *pallidum* genetic population structure and relevance to syphilis prevention and treatment"

**Supplementary Materials and Methods**

**Ethics statements**

This study was reviewed and approved by the institutional review board (IRB) at the University of North Carolina at Chapel Hill (protocol number 19-0311) and University of Washington (protocol STUDY00000885). Each clinical site also obtained approval from their respective local IRBs before beginning recruitment and enrollment. Additional IRB approvals and protocol numbers are as follows: China, Southern Medical University (protocol GDDHLS-20181202 [R3]); Colombia, Centro Internacional de Entrenamiento e Investigaciones Medicas, CIDEIM IHREC IRB (protocol 1289); Malawi, National Health Sciences Research Committee, Ministry of Health and Population (protocol 2252; Argentina, Ethical committee, Hospital de Niños Dr. Ricardo Gutierrez (protocol 22.07); Vietnam, UNC (protocol 21-3181) and National Hospital of Dermatology and Venereology (protocol 214/HDDD-BVDLTW); Sri Lanka, ERC of Medical Faculty, University of Colombo (protocol EC-22-017); Japan, National Institute of Infectious Diseases (protocols 508 and 705); Peru, University of Southern California (protocol HS-21-00353); Madagascar, University of Alabama at Birmingham Institutional Review Board for Human Subjects; Brazil, National Research Ethics Commission (protocol 56591822.9.0000.5243).

All clinical research activities adhered to the Declaration of Helsinki guidelines. Written informed consent was obtained from all participants prior to enrollment. All samples were de-identified after collection.

**Samples and whole-genome sequencing**

Cross-sectional samples were collected from patients with early syphilis (primary, secondary, early latent syphilis) from multiple sites in Argentina, China, Colombia, Malawi, Vietnam, and Sri Lanka. We also sequenced banked samples collected as part of separate studies in Brazil, China, Ireland, Japan, Madagascar, and Peru.

Samples were extracted from lesion swabs, skin biopsies, and whole blood samples as previously described [(1,2)](https://sciwheel.com/work/citation?ids=16514573,17000479&pre=&pre=&suf=&suf=&sa=0,0). *T. pallidum* sequencing was performed at the University of North Carolina at Chapel Hill (UNC) and the University of Washington (UW). *T. pallidum* burdens were quantified using quantitative real-time polymerase chain reaction (qPCR) targeting the *polA* (*tp0105*) [(2)](https://sciwheel.com/work/citation?ids=17000479&pre=&suf=&sa=0) or *tp47* (*tp0574*) gene multiplex with human β-globin [(3)](https://sciwheel.com/work/citation?ids=16514565&pre=&suf=&sa=0). Samples were selected for *T. pallidum* whole-genome enrichment using oligonucleotide probes (RNA baits by Agilent Technologies, Santa Clara, CA, at UNC; DNA baits by IDT DNA Technologies at UW), and sequenced with paired-end 150 bp reads using MiSeq at UNC (Illumina, San Diego, CA) and paired-end 151 bp reads using NextSeq2000 or NovaSeq6000 (Illumina) at UW, as previously described [(2,4)](https://sciwheel.com/work/citation?ids=17000479,15460703&pre=&pre=&suf=&suf=&sa=0,0).

Sequence reads underwent quality control using FastQC (v0.12.1) [(5)](https://sciwheel.com/work/citation?ids=16514624&pre=&suf=&sa=0). Adapters were trimmed with Trimmomatic (v0.39) [(6)](https://sciwheel.com/work/citation?ids=63413&pre=&suf=&sa=0). Host genome sequences were identified and removed using bbmap (v38.82) [(7)](https://sciwheel.com/work/citation?ids=16514625&pre=&suf=&sa=0), by mapping sequences to human (hg19) and rabbit (oryCun2) genomes. Potential bacterial contamination was assessed using StrainSeeker (v1.5.1) [(8)](https://sciwheel.com/work/citation?ids=3644280&pre=&suf=&sa=0). Sequence alignment to reference genomes (Nichols CP004010.2 and SS14 CP004011.1) was performed using bwa (v0.7.17) [(9)](https://sciwheel.com/work/citation?ids=12643106&pre=&suf=&sa=0). Post-alignment filtering included the removal of duplicate reads, indel realignment, and the exclusion of reads with excessive mismatches, soft and hard clips, chimeric alignments, repetitively aligned reads, and low-quality mappings, as detailed previously [(2)](https://sciwheel.com/work/citation?ids=17000479&pre=&suf=&sa=0). Genomic sequences with at least three unique reads covering over 80% of the genome were retained for analysis. Variant calling was conducted using GATK (v4.4) [(10)](https://sciwheel.com/work/citation?ids=16514626&pre=&suf=&sa=0).

We combined our data with publicly available genomic sequencing data from 1409 samples downloaded from the Sequence Read Archive (SRA Supplementary Table 1). All downloaded raw data were subjected to the same variant calling procedure.

Macrolide resistance-associated mutations (A2058G and A2059G) were identified using a competitive mapping approach against a reference 23S rRNA alignment. Variant calling was performed using both GATK and Samtools (v1.17) [(11)](https://sciwheel.com/work/citation?ids=10500701&pre=&suf=&sa=0) and, in cases of discrepancy between the two callers, aligned reads were manually inspected to resolve the presence or absence of resistance mutations.

**Targeted long-read sequencing of the *tpr* family**

Genomic regions corresponding to the *T. pallidum* *tpr* gene family subfamilies I and II, namely *tp0117* (*tprC*), *tp0131* (*tprD*), *tp0313* (*tprE*), *tp0316* (*tprF*), *tp0317* (*tprG*), *tp0620* (*tprI*), and *tp0621* (*tprJ*), were sequenced from 206 clinical strains collected in China, Vietnam, Colombia, and Malawi. All sequencing procedures were carried out according to a modified protocol described by Pospíšilová et al. [(12)](https://sciwheel.com/work/citation?ids=18362043&pre=&suf=&sa=0). DNA was extracted using the DNeasy Blood and Tissue Kit (QIAGEN, 69506), followed by whole-genome amplification with the REPLI-g kit (QIAGEN) in three pooled parallel reactions. Nested PCR targeting the regions of interest was performed using PrimeSTAR GXL DNA Polymerase (Takara Bio), employing a touchdown PCR scheme: initial denaturation at 94°C for 1 min; eight cycles of 98°C for 10 s, annealing at 68°C for 15 s (decreasing by 1°C per cycle), and extension at 68°C for 1 min 45 s; 35 cycles of 98°C for 10 s, 61°C for 15 s, and 68°C for 6 min; and a final extension at 68°C for 7 min. Outer and inner PCR products were amplified under identical conditions.

Amplicons were purified by SPRI bead cleanup (AMPure XP Beads, Beckman Coulter, A63881) and pooled equimolarly for long-read sequencing on the Oxford Nanopore MinION platform. Library preparation used the Native Barcoding Kit 96 v14 (Oxford Nanopore Technologies, SQK-NBD114.96) with minor modifications, including extended incubation and EB buffer resuspension at 37°C. Sequencing was performed on R10.4.1 flow cells (FLO-MIN114) for 48 h.

Basecalling and demultiplexing were conducted using Guppy v4.4.1 (high-accuracy model; Q20 threshold) . Cutadapt [(13)](https://sciwheel.com/work/citation?ids=827836&pre=&suf=&sa=0) was used for adapter trimming, and reads were mapped to masked reference genomes (CP004010.2 and CP004011.1) with minimap2 [(14)](https://sciwheel.com/work/citation?ids=5243528&pre=&suf=&sa=0). Aligned reads were processed with SAMtools [(15)](https://sciwheel.com/work/citation?ids=48787&pre=&suf=&sa=0), NGSUtils [(16)](https://sciwheel.com/work/citation?ids=235798&pre=&suf=&sa=0), and Picard (<http://broadinstitute.github.io/picard/>); reads with more than 20% soft clipping or greater than 10% mismatch rates were excluded. Reads of 1–6 kb and spanning the regions of interest ±500 nt were retained. Consensus sequences were assembled with Canu v1.8 [(17)](https://sciwheel.com/work/citation?ids=3748382&pre=&suf=&sa=0) and polished using Racon v1.4.13 [(18)](https://sciwheel.com/work/citation?ids=3979196&pre=&suf=&sa=0) and Medaka v0.7.1 ([https://github.com/nanoporetech/medaka](https://github.com/nanoporetech/medaka)%22.%5B3)).

**Population analysis**

For WGS analyses, regions that are highly repetitive and difficult to sequence, namely the *tpr* gene family, *arp*, *tp0470*, and 23S rRNA, were manually masked in all Nichols- and SS14-lineage genomes prior to downstream analysis. Only single nucleotide polymorphisms (SNPs) were retained for population structure and phylogeny-based analyses. We used FastBAPS (v1.0.8) to estimate the number of distinct *T. pallidum* subpopulations within each *T. pallidum* lineage. Multiple sequence alignment was performed using MAFFT (v7.490) [(19)](https://sciwheel.com/work/citation?ids=387873&pre=&suf=&sa=0). Population structure was further assessed using principal component analysis (PCA) via the EIGENSTRAT tool from EIGENSOFT (v6.1) [(20)](https://sciwheel.com/work/citation?ids=431745&pre=&suf=&sa=0). We calculated Weir and Cockerham F-Statistics (Fst) [(21)](https://sciwheel.com/work/citation?ids=30216&pre=&suf=&sa=0) with VCFtools [(22)](https://sciwheel.com/work/citation?ids=111675&pre=&suf=&sa=0) identifying SNPs with Fst > 0.9 as significantly differentiated. SNP functional annotation was performed using SnpEff (v5.1) [(23)](https://sciwheel.com/work/citation?ids=511142&pre=&suf=&sa=0).

Whole-genome phylogenies were constructed using IQ-TREE 2 [(24)](https://sciwheel.com/work/citation?ids=8188277&pre=&suf=&sa=0) with the GTR+F+I+G nucleotide substitution model and 10,000 ultrafast bootstrap replicates. Phylogenetic trees were visualized and annotated using the ggtree package [(25)](https://sciwheel.com/work/citation?ids=2006241&pre=&suf=&sa=0) in R (v4.3.1) [(26)](https://sciwheel.com/work/citation?ids=16516445&pre=&suf=&sa=0). GrapeTree [(27)](https://sciwheel.com/work/citation?ids=5654265&pre=&suf=&sa=0) was used to generate minimum spanning trees of selected genes. Sequence types (STs) were assigned using the MLST scheme described by Grillová et al. [(28)](https://sciwheel.com/work/citation?ids=12179350&pre=&suf=&sa=0), and the PubMLST.org database [(29)](https://sciwheel.com/work/citation?ids=5981516&pre=&suf=&sa=0), utilizing MLST software (v2.16.1). Visualizations and plots were produced using R and the ggplot2 package (v3.4.3) [(30)](https://sciwheel.com/work/citation?ids=16516453&pre=&suf=&sa=0).

**Supplementary References**

[1.	Lieberman NAP, Armstrong TD, Chung B, Pfalmer D, Hennelly CM, Haynes A, et al. High-throughput nanopore sequencing of Treponema pallidum tandem repeat genes arp and tp0470 reveals clade-specific patterns and recapitulates global whole genome phylogeny. Front Microbiol. 2022 Sep 20;13:1007056.](https://sciwheel.com/work/bibliography/16514573)

[2.	Seña AC, Matoga MM, Yang L, Lopez-Medina E, Aghakhanian F, Chen JS, et al. Clinical and genomic diversity of Treponema pallidum subspecies pallidum to inform vaccine research: an international, molecular epidemiology study. Lancet Microbe. 2024 Sep;5(9):100871.](https://sciwheel.com/work/bibliography/17000479)

[3.	Lieberman NAP, Lin MJ, Xie H, Shrestha L, Nguyen T, Huang M-L, et al. Treponema pallidum genome sequencing from six continents reveals variability in vaccine candidate genes and dominance of Nichols clade strains in Madagascar. PLoS Negl Trop Dis. 2021 Dec 22;15(12):e0010063.](https://sciwheel.com/work/bibliography/16514565)

[4.	Lieberman NAP, Avendaño CC, Bakhash SAKM, Nunley E, Xie H, Giacani L, et al. Genomic Epidemiology of Treponema pallidum and Circulation of Strains With Diminished tprK Antigen Variation Capability in Seattle, 2021-2022. J Infect Dis. 2024 Mar 14;229(3):866–75.](https://sciwheel.com/work/bibliography/15460703)

[5.	Andrews S. FastQC: a quality control tool for high throughput sequence data. [Internet]. 2010. Available from: http://www.bioinformatics.babraham.ac.uk/projects/fastqc](https://sciwheel.com/work/bibliography/16514624)

[6.	Bolger AM, Lohse M, Usadel B. Trimmomatic: A flexible trimmer for Illumina sequence data. Bioinformatics. 2014 Aug 1;30(15):2114–20.](https://sciwheel.com/work/bibliography/63413)

[7.	Bushnell B. BBMap:  A Fast, Accurate, Splice-Aware Aligner. Report Number: LBNL-7065E. Research Org.: Lawrence Berkeley National Lab. (LBNL), Berkeley, CA (United States); 2014.](https://sciwheel.com/work/bibliography/16514625)

[8.	Roosaare M, Vaher M, Kaplinski L, Möls M, Andreson R, Lepamets M, et al. StrainSeeker: fast identification of bacterial strains from raw sequencing reads using user-provided guide trees. PeerJ. 2017 May 18;5:e3353.](https://sciwheel.com/work/bibliography/3644280)

[9.	Li H. Aligning sequence reads, clone sequences and assembly contigs with BWA-MEM. arXiv. 2013;](https://sciwheel.com/work/bibliography/12643106)

[10.	Van der Auwera G, O’Connor B. Genomics in the Cloud: Using Docker, GATK, and WDL in Terra. 1st ed. Sebastopol, CA: O’Reilly Media; 2020.](https://sciwheel.com/work/bibliography/16514626)

[11.	Danecek P, Bonfield JK, Liddle J, Marshall J, Ohan V, Pollard MO, et al. Twelve years of SAMtools and BCFtools. Gigascience. 2021 Feb 16;10(2).](https://sciwheel.com/work/bibliography/10500701)

[12.	Pospíšilová P, Fedrová P, Vrbová E, Hennelly CM, Aghakhanian F, Hawley KL, et al. Analysis of Treponema pallidum subsp. pallidum predicted outer membrane proteins (OMPeomes) in 21 clinical samples: variant sequences are predominantly surface-exposed. mSphere. 2025 Sep 30;10(9):e0021325.](https://sciwheel.com/work/bibliography/18362043)

[13.	Martin M. Cutadapt removes adapter sequences from high-throughput sequencing reads. EMBnet j. 2011 May 2;17(1):10.](https://sciwheel.com/work/bibliography/827836)

[14.	Li H. Minimap2: pairwise alignment for nucleotide sequences. Bioinformatics. 2018 Sep 15;34(18):3094–100.](https://sciwheel.com/work/bibliography/5243528)

[15.	Li H, Handsaker B, Wysoker A, Fennell T, Ruan J, Homer N, et al. The Sequence Alignment/Map format and SAMtools. Bioinformatics. 2009 Aug 15;25(16):2078–9.](https://sciwheel.com/work/bibliography/48787)

[16.	Breese MR, Liu Y. NGSUtils: a software suite for analyzing and manipulating next-generation sequencing datasets. Bioinformatics. 2013 Feb 15;29(4):494–6.](https://sciwheel.com/work/bibliography/235798)

[17.	Koren S, Walenz BP, Berlin K, Miller JR, Bergman NH, Phillippy AM. Canu: scalable and accurate long-read assembly via adaptive k-mer weighting and repeat separation. Genome Res. 2017 May;27(5):722–36.](https://sciwheel.com/work/bibliography/3748382)

[18.	Vaser R, Sović I, Nagarajan N, Šikić M. Fast and accurate *de novo* genome assembly from long uncorrected reads. Genome Res. 2017 May;27(5):737–46.](https://sciwheel.com/work/bibliography/3979196)

[19.	Katoh K, Standley DM. MAFFT multiple sequence alignment software version 7: improvements in performance and usability. Mol Biol Evol. 2013 Apr;30(4):772–80.](https://sciwheel.com/work/bibliography/387873)

[20.	Price AL, Patterson NJ, Plenge RM, Weinblatt ME, Shadick NA, Reich D. Principal components analysis corrects for stratification in genome-wide association studies. Nat Genet. 2006 Aug;38(8):904–9.](https://sciwheel.com/work/bibliography/431745)

[21.	Weir BS, Cockerham CC. Estimating F-Statistics for the Analysis of Population Structure. Evolution. 1984 Nov 1;38(6):1358.](https://sciwheel.com/work/bibliography/30216)

[22.	Danecek P, Auton A, Abecasis G, Albers CA, Banks E, DePristo MA, et al. The variant call format and VCFtools. Bioinformatics. 2011 Aug 1;27(15):2156–8.](https://sciwheel.com/work/bibliography/111675)

[23.	Cingolani P, Platts A, Wang LL, Coon M, Nguyen T, Wang L, et al. A program for annotating and predicting the effects of single nucleotide polymorphisms, SnpEff: SNPs in the genome of *Drosophila melanogaster* strain w1118; iso-2; iso-3. Fly (Austin). 2012 Jun;6(2):80–92.](https://sciwheel.com/work/bibliography/511142)

[24.	Minh BQ, Schmidt HA, Chernomor O, Schrempf D, Woodhams MD, von Haeseler A, et al. IQ-TREE 2: New models and efficient methods for phylogenetic inference in the genomic era. Mol Biol Evol. 2020 May 1;37(5):1530–4.](https://sciwheel.com/work/bibliography/8188277)

[25.	Yu G, Smith DK, Zhu H, Guan Y, Lam TTY. ggtree: An R package for visualization and annotation of phylogenetic trees with their covariates and other associated data. Methods Ecol Evol. 2016 Aug;8:28–36.](https://sciwheel.com/work/bibliography/2006241)

[26.	R Core Team. R: A Language and Environment for Statistical Computing. Vienna, Austria: R Foundation for Statistical Computing; 2023.](https://sciwheel.com/work/bibliography/16516445)

[27.	Zhou Z, Alikhan N-F, Sergeant MJ, Luhmann N, Vaz C, Francisco AP, et al. GrapeTree: visualization of core genomic relationships among 100,000 bacterial pathogens. Genome Res. 2018 Sep;28(9):1395–404.](https://sciwheel.com/work/bibliography/5654265)

[28.	Grillová L, Bawa T, Mikalová L, Gayet-Ageron A, Nieselt K, Strouhal M, et al. Molecular characterization of Treponema pallidum subsp. pallidum in Switzerland and France with a new multilocus sequence typing scheme. PLoS ONE. 2018 Jul 30;13(7):e0200773.](https://sciwheel.com/work/bibliography/12179350)

[29.	Jolley KA, Bray JE, Maiden MCJ. Open-access bacterial population genomics: BIGSdb software, the PubMLST.org website and their applications. [version 1; peer review: 2 approved]. Wellcome Open Res. 2018 Sep 24;3:124.](https://sciwheel.com/work/bibliography/5981516)

[30.	Wickham H. ggplot2: Elegant Graphics for Data Analysis. 2nd ed. Cham: Springer; 2016.](https://sciwheel.com/work/bibliography/16516453)

**Supplementary Figures**

**Supplementary Figure 1:** Overview of participants and genomes in this study, detailing the number of sequences per country, patient age at sample collection (where available), *Treponema pallidum* subsp. *pallidum* (TPA) sublineage ratio by country, disease stage, and patient sex and sexuality.

**Supplementary Figure 2:** Maximum likelihood phylogeny of TPA genomes from this study as well as the Nichols, SS14, Mexico-A TPA reference genomes and the Samoa D *Treponema pallidum* subsp. *pertenue* (TPE) genome. Annotations shown in gray indicate unknown attributes.

**Supplementary Figure 3:** Maximum likelihood phylogeny of Mexico-A-like strains, including SS14 and Nichols reference TPA genomes and the Samoa_D TPE genome.

**Supplementary Figure 4:** Minimum spanning tree (MST) of new genomes generated during this study (n=298) by lineage: (A) Nichols and (B) SS14. Trees are color-coded by country of origin.

**Supplementary Figure 5:** Minimum spanning tree (MST) of both genomes generated in this study and publicly available genomes (n=1,707 total) by lineage: (A) Nichols and (B) SS14. Trees are color-coded by country of origin.

**Supplementary Figure 6:** Principal component analysis (PCA) of Nichols-lineage genomes from Madagascar, Malawi, Tanzania, and South Africa.

**Supplementary Figure 7:** Identification of SNPs that differentiate selected TPA subpopulations. A) Fixation index (F_st_) analysis comparing the Nichols-4 subpopulation (primarily genomes from South America and Oceania) and Nichols-5 and -6 (genomes from Africa) shows an accumulation of non-synonymous SNPs in *tp0865* and *tp0462*. Minimum spanning trees (MSTs) of global sequences illustrate greater diversity of these genes in South America and Oceania. B) F_st_ analysis comparing the two African Nichols-lineage subpopulations, Nichols-5 and -6, shows an accumulation of non-synonymous SNPs in *tp0136* and *tp0483*. MSTs of global sequences show higher diversity in *tp0136* and *tp0483* in TPA from Africa compared to other continents. C) F_st_ analysis of SS14-lineage subpopulations SS14-4 and -5, both of which appear as extremes along PC1 in the PCA, reveals the presence of non-synonymous SNPs in *TP0705* (*mrcA*).

**Supplementary Figure 8:** Minimum spanning tree (MST) of new genomes generated during this study in the (A) Nichols and (B) SS14 lineages, annotated by sequence type (ST) determined by MLST. Abbreviations: ND, strain type not determined by MLST.

**Supplementary Figure 9:** Minimum spanning tree (MST) restricted to sequences of non-*tpr* OMP genes including OMP assembly machinery (BamA and LptD), long-chain-fatty-acid transporters (FadLs), 8-stranded β-barrels and OM factors (OMFs) for efflux pumps (Supplementary Table 3). The tree is color-coded by Nichols and SS14 subpopulations assigned by FastBAPS using whole-genome sequencing data.

**Supplementary Figure 10:** A) Frequency of macrolide resistance markers in the TPA 23S rRNA gene among strains sequenced in this study. B) Frequency of the *tp0705* (*mrca*) A1873G mutation recently associated with ceftriaxone/penicillin G tolerance in vitro.

**Supplementary Figure 11:** Minimum spanning tree (MST) depicting genetic relationships within *tpr* subfamily I (*tprC, tprD*, and *tprI*), color-coded by subpopulation.

**Supplementary Figure 12:** Minimum spanning tree (MST) depicting genetic relationships within *tpr* subfamily I (*tprE*, *tprG*, and *tprJ*), color-coded by subpopulation.

**Supplementary Figure 1:** Overview of participants and genomes in this study, detailing the number of sequences per country, patient age at sample collection (where available), *Treponema pallidum* subsp. *pallidum* (TPA) sublineage ratio by country, disease stage, and patient sex and sexuality.

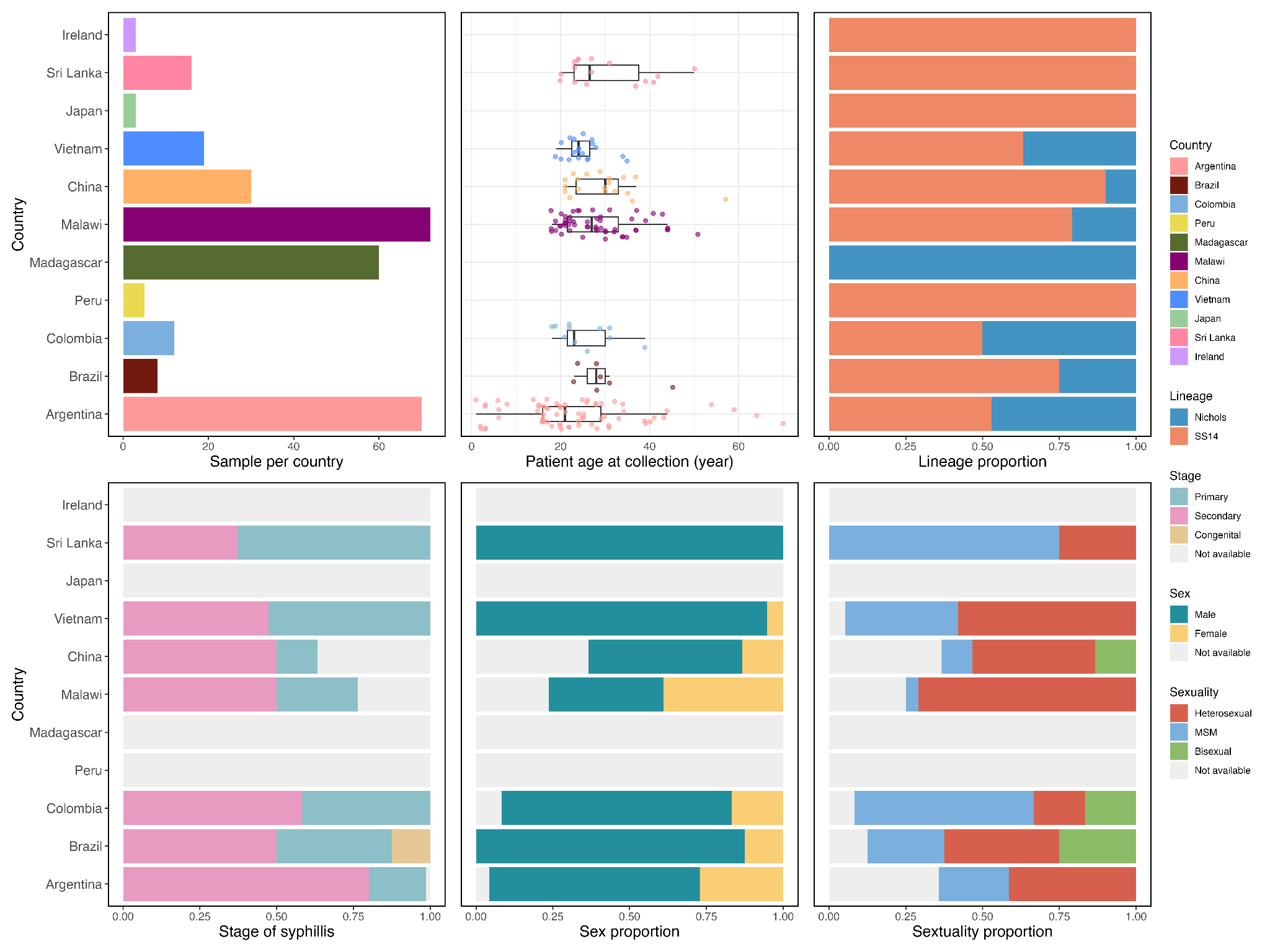

**Supplementary Figure 2:** Maximum likelihood phylogeny of TPA genomes from this study as well as the Nichols, SS14, Mexico-A TPA reference genomes and the Samoa_D *Treponema pallidum* subsp. *pertenue* (TPE) genome. Annotations shown in gray indicate unknown attributes.
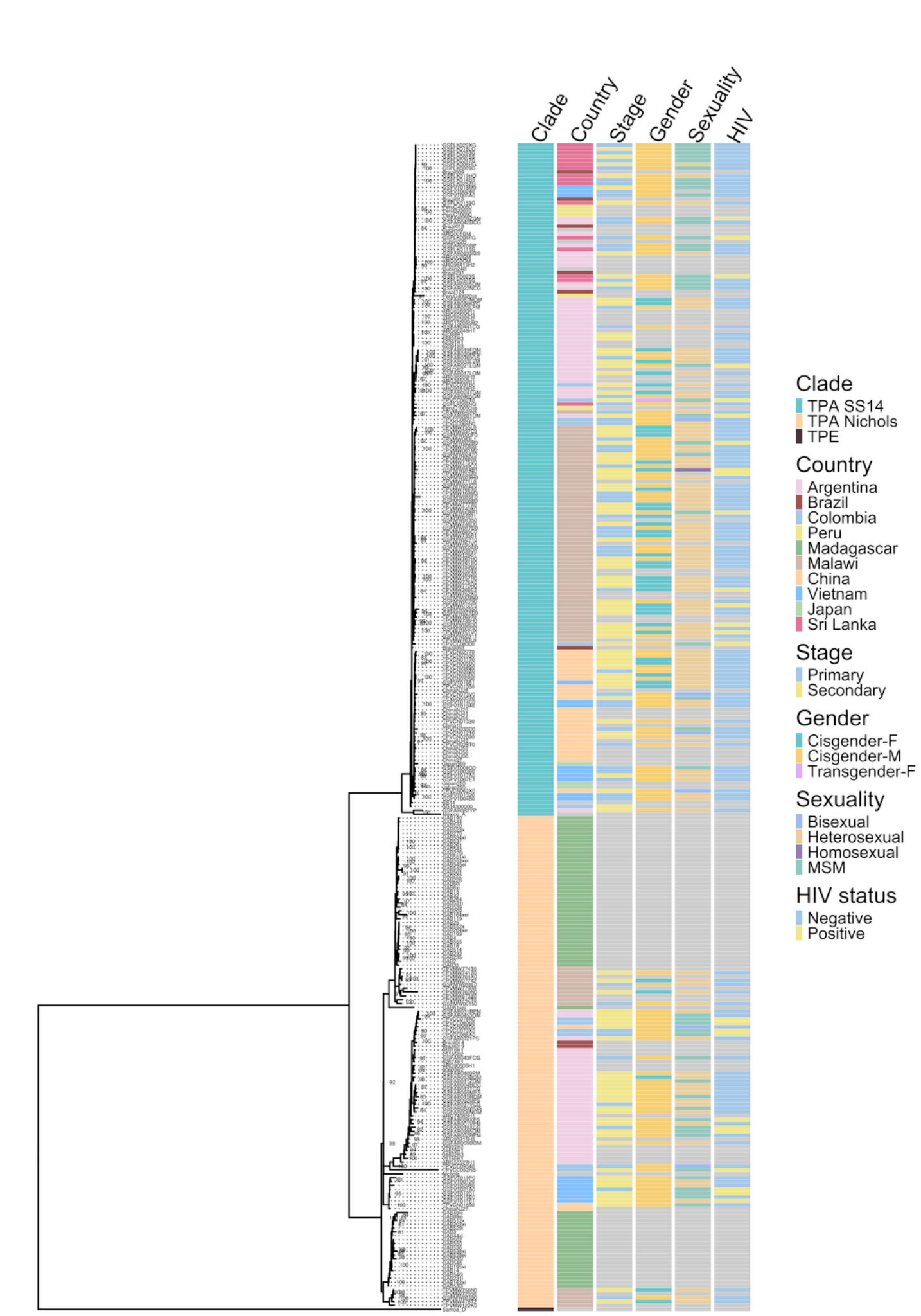

**Supplementary Figure 3:** Maximum likelihood phylogeny of Mexico-A-like strains, including SS14 and Nichols reference TPA genomes and the Samoa_D TPE genome.

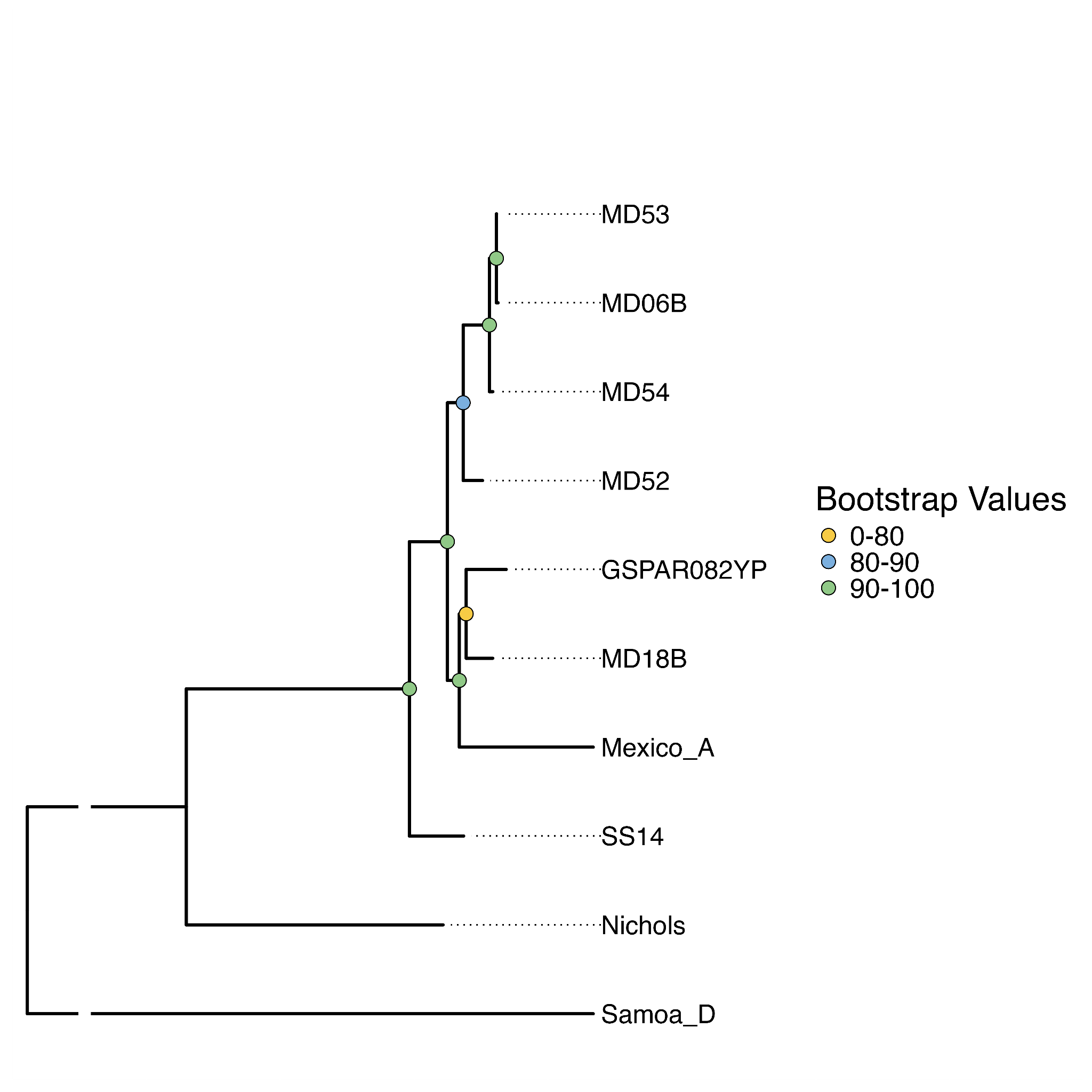

**Supplementary Figure 4:** Minimum spanning tree (MST) of new genomes generated during this study (n=298) by lineage: (A) Nichols and (B) SS14. Trees are color-coded by country of origin.

A. Nichols lineage

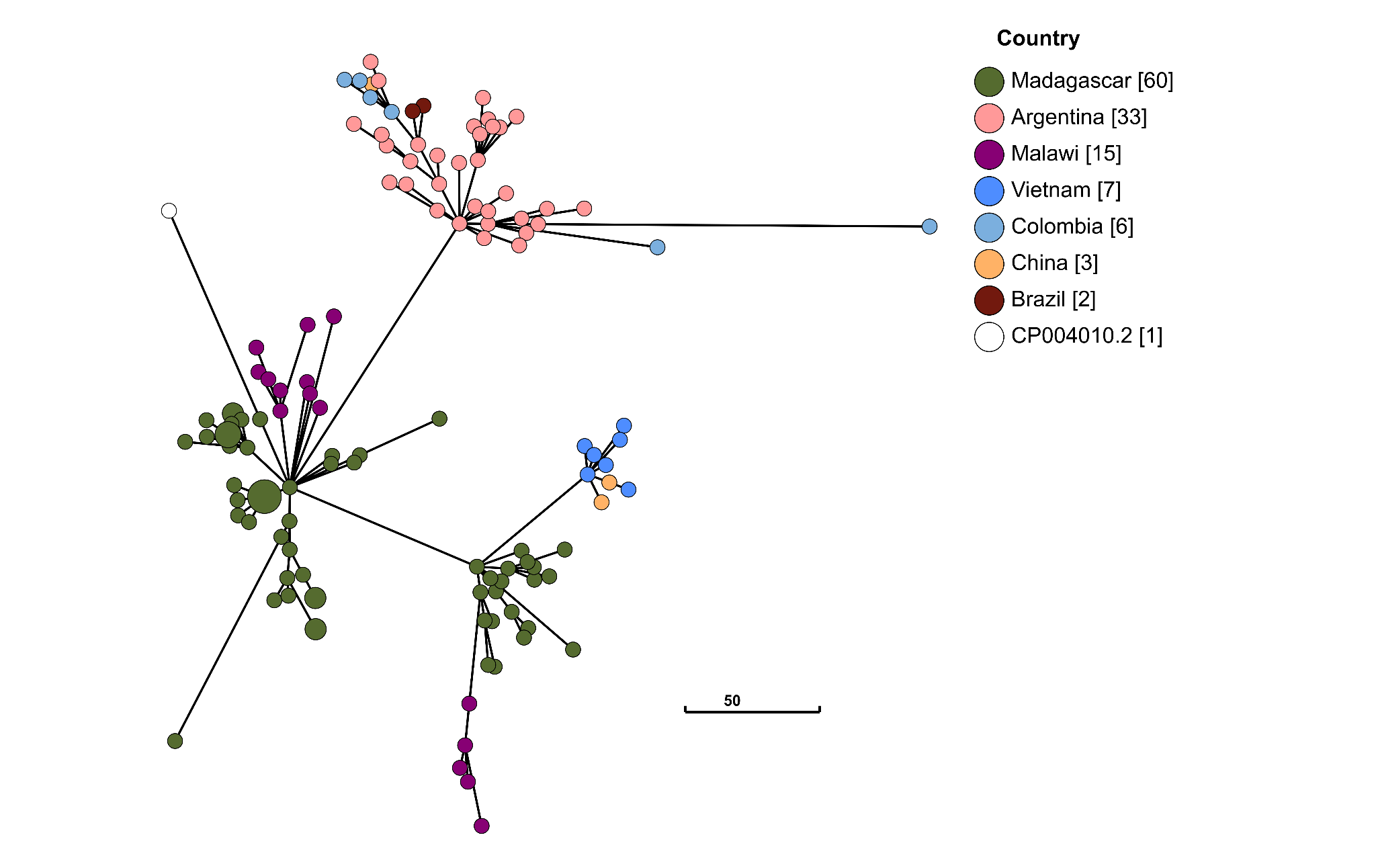

B. SS14 lineage

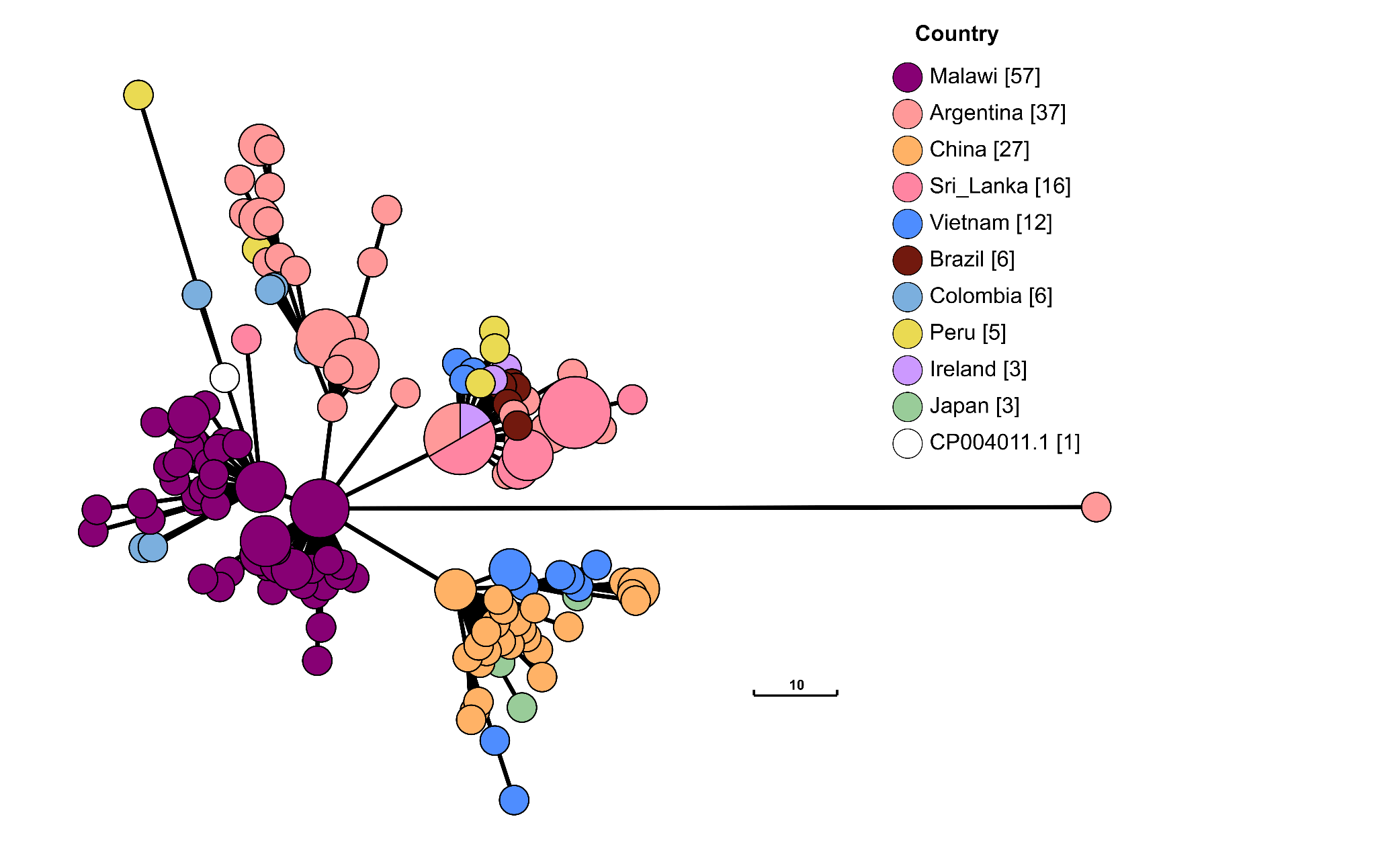

**Supplementary Figure 5:** Minimum spanning tree (MST) of both genomes generated in this study and publicly available genomes (n=1,707 total) by lineage: (A) Nichols and (B) SS14. Trees are color-coded by country of origin.

A. Nichols lineage

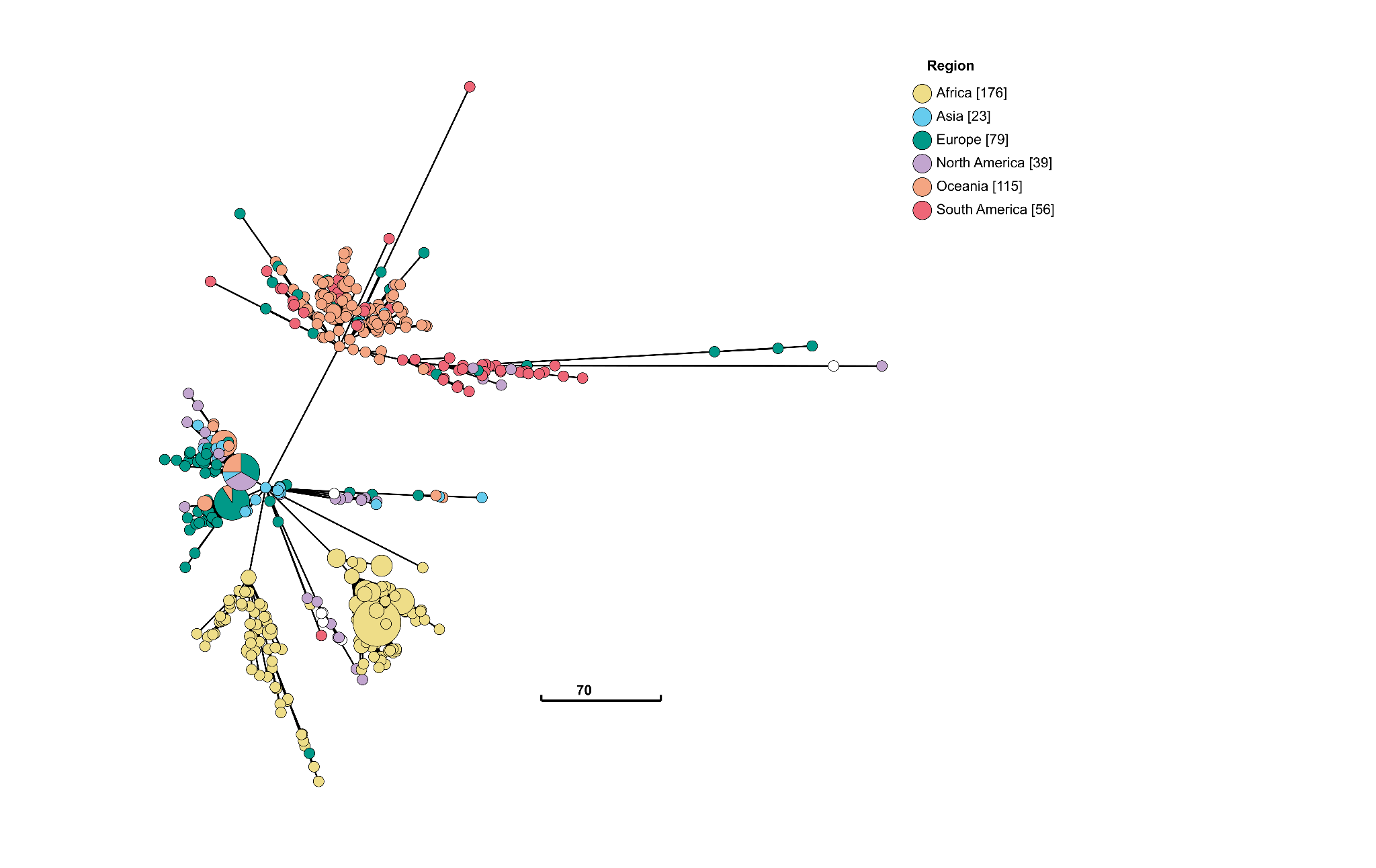

B. SS14 lineage

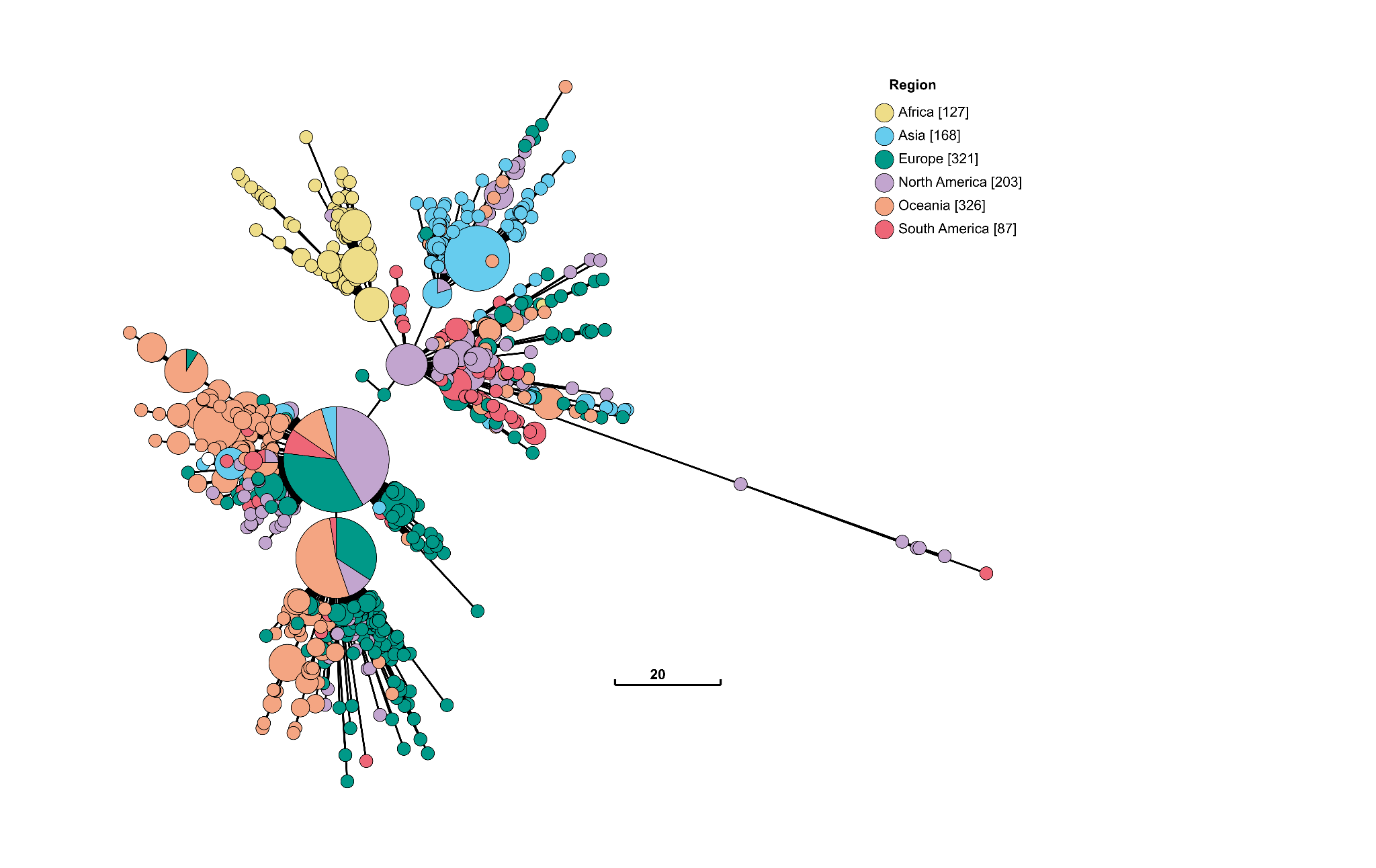

**Supplementary Figure 6:** Principal component analysis (PCA) of Nichols-lineage genomes from Madagascar, Malawi, Tanzania, and South Africa.

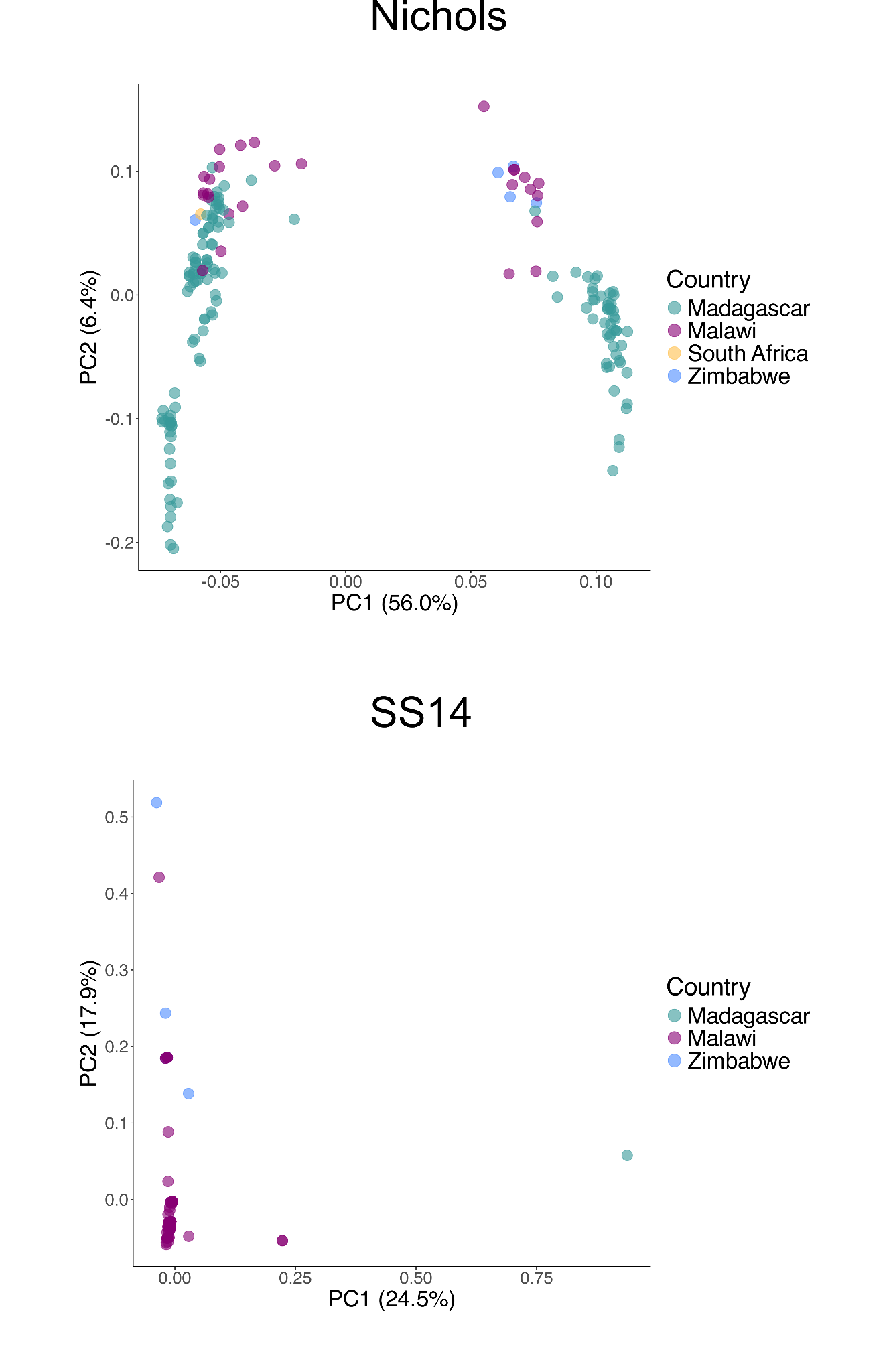

**Supplementary Figure 7:** Identification of SNPs that differentiate selected TPA subpopulations. A) Fixation index (F_st_) analysis comparing the Nichols-4 subpopulation (primarily genomes from South America and Oceania) and Nichols-5 and -6 (genomes from Africa) shows an accumulation of non-synonymous SNPs in *tp0865* and *tp0462*. Minimum spanning trees (MSTs) of global sequences illustrate greater diversity of these genes in South America and Oceania. B) F_st_ analysis comparing the two African Nichols-lineage subpopulations, Nichols-5 and -6, shows an accumulation of non-synonymous SNPs in *tp0136* and *tp0483*. MSTs of global sequences show higher diversity in *tp0136* and *tp0483* in TPA from Africa compared to other continents. C) F_st_ analysis of SS14-lineage subpopulations SS14-4 and -5, both of which appear as extremes along PC1 in the PCA, reveals the presence of non-synonymous SNPs in *TP0705* (*mrcA*).

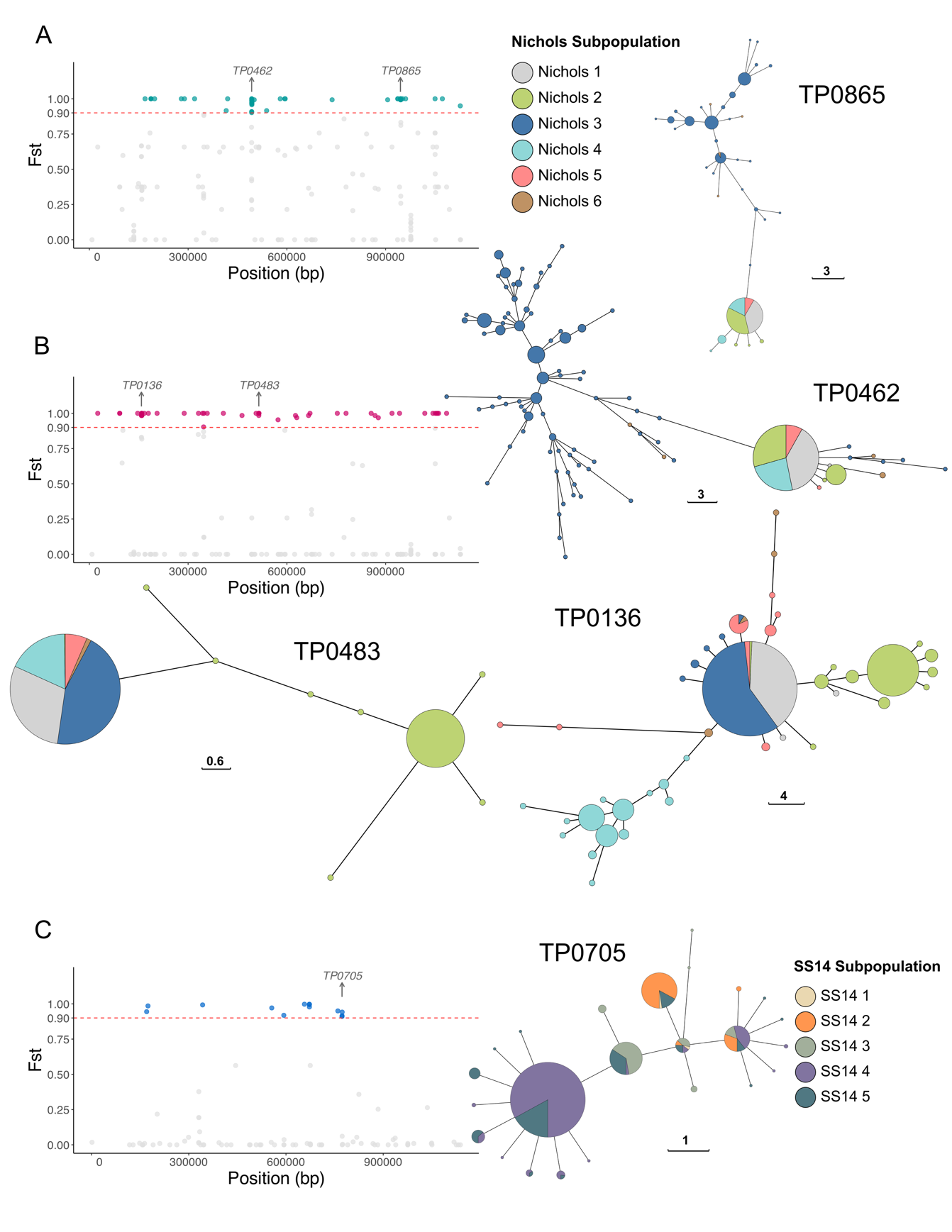

**Supplementary Figure 8:** Minimum spanning tree (MST) of new genomes generated during this study in the (A) Nichols and (B) SS14 lineages, annotated by sequence type (ST) determined by MLST. Abbreviations: ND, strain type not determined by MLST.

A) Nichols lineage

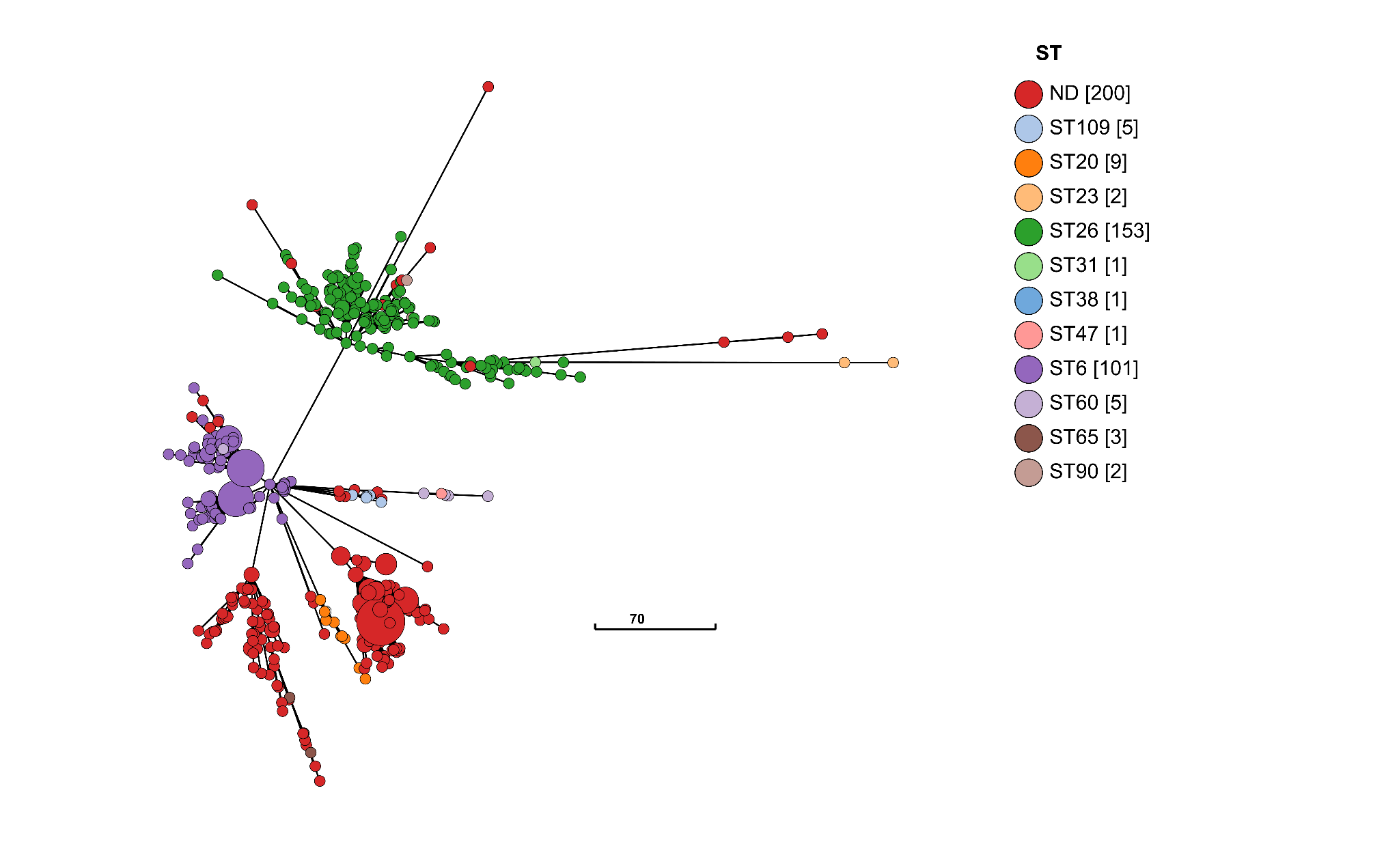

B) SS14 lineage

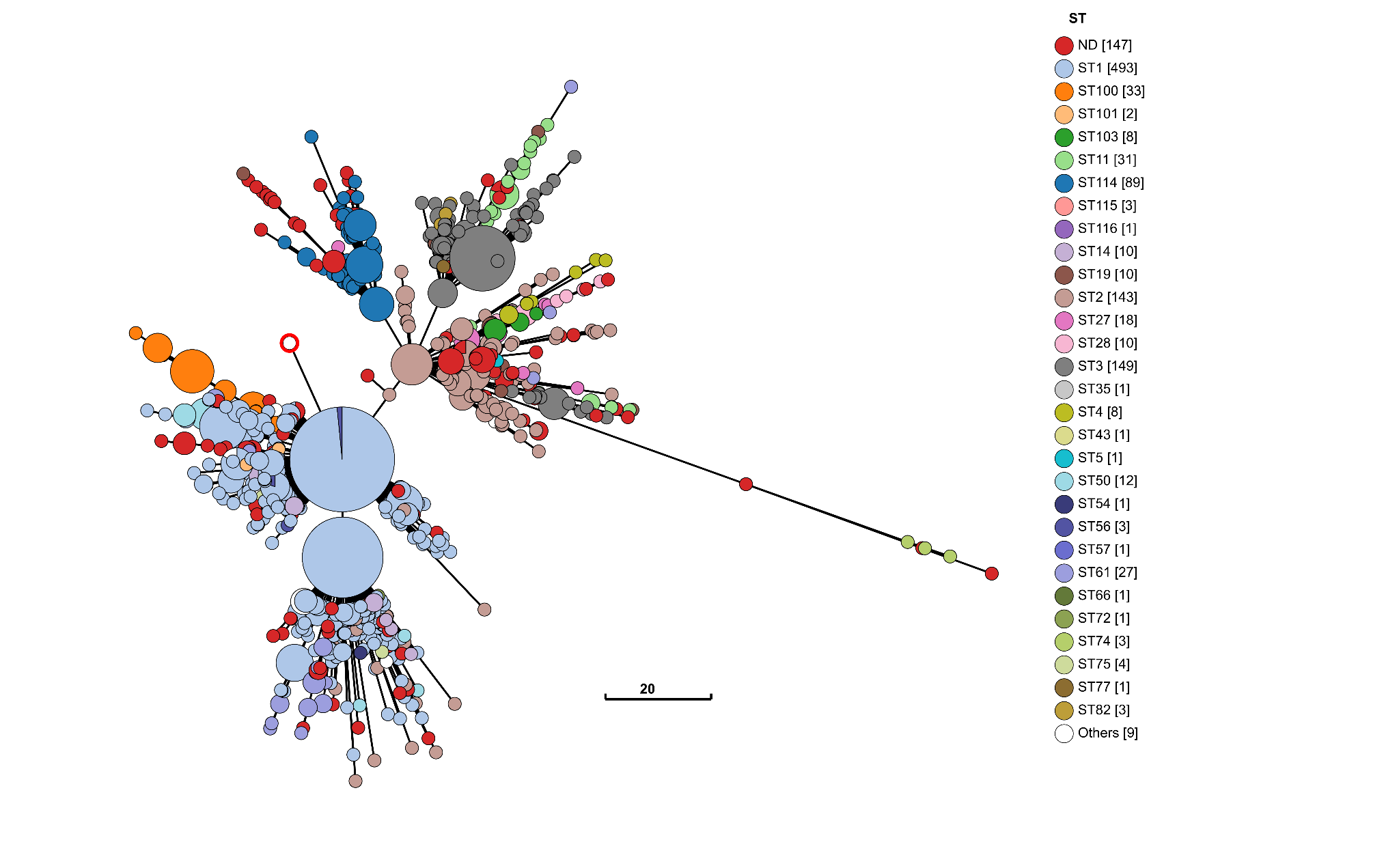

**Supplementary Figure 9:** Minimum spanning tree (MST) restricted to sequences of non-*tpr* OMP genes. The tree is color-coded by Nichols and SS14 subpopulations assigned by FastBAPS using whole-genome sequencing data.

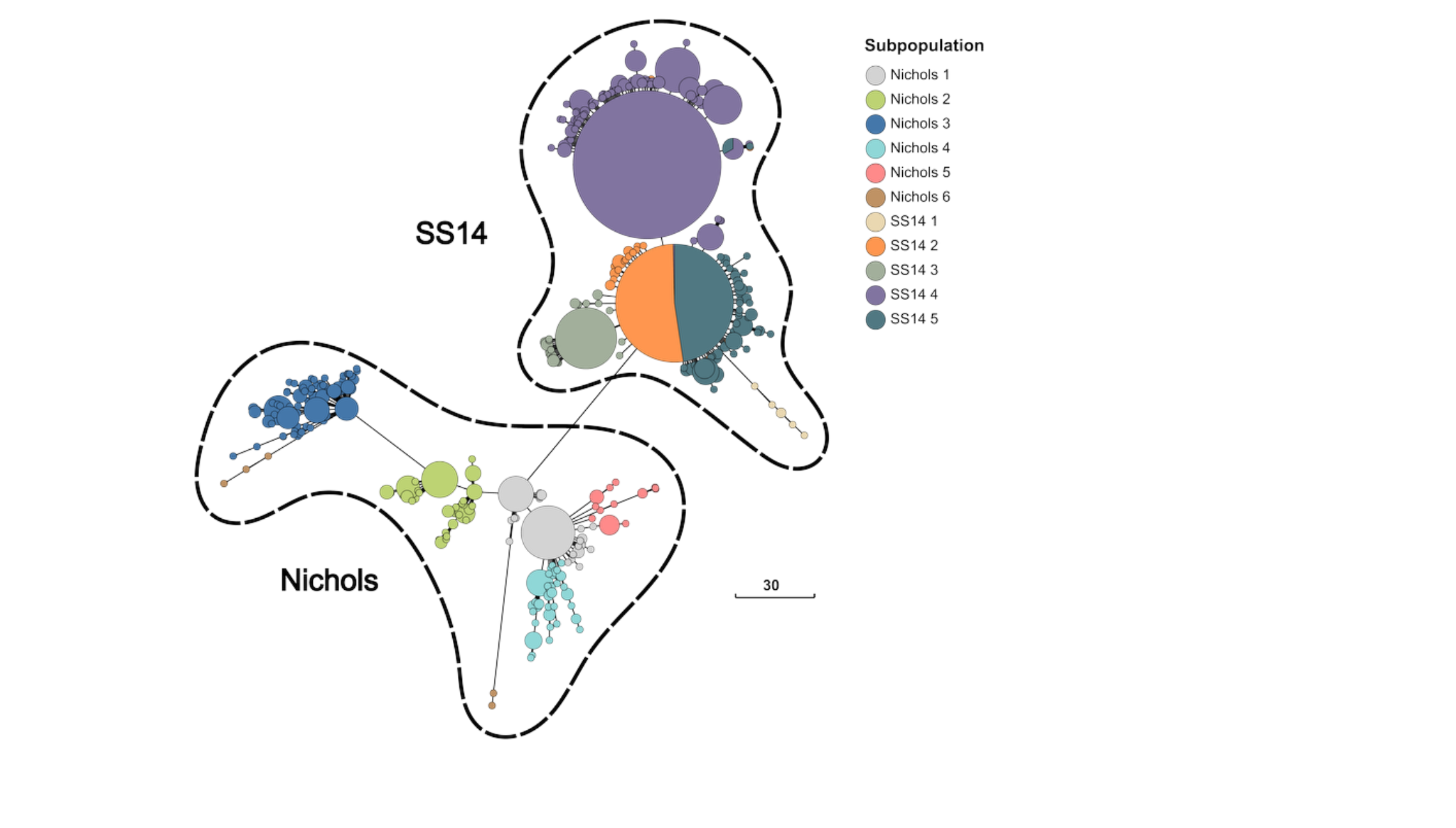

**Supplementary Figure 10:** Minimum spanning tree (MST) depicting genetic relationships within *tpr* subfamily I (*tprC, tprD*, and *tprI*), color-coded by subpopulation.

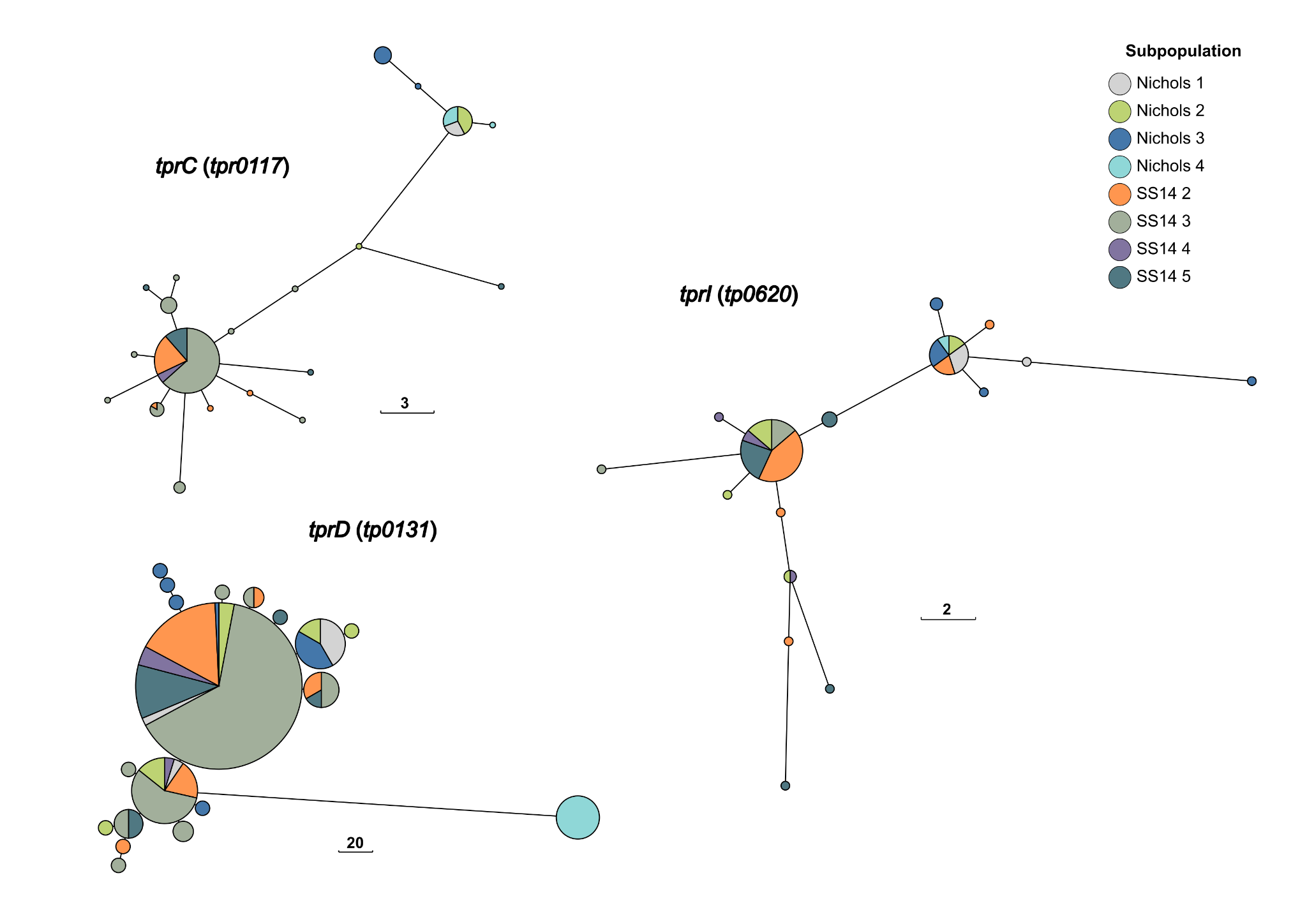

**Supplementary Figure 11:** Minimum spanning tree (MST) depicting genetic relationships within *tpr* subfamily I (*tprE, tprG*, and *tprJ*), color-coded by subpopulation.

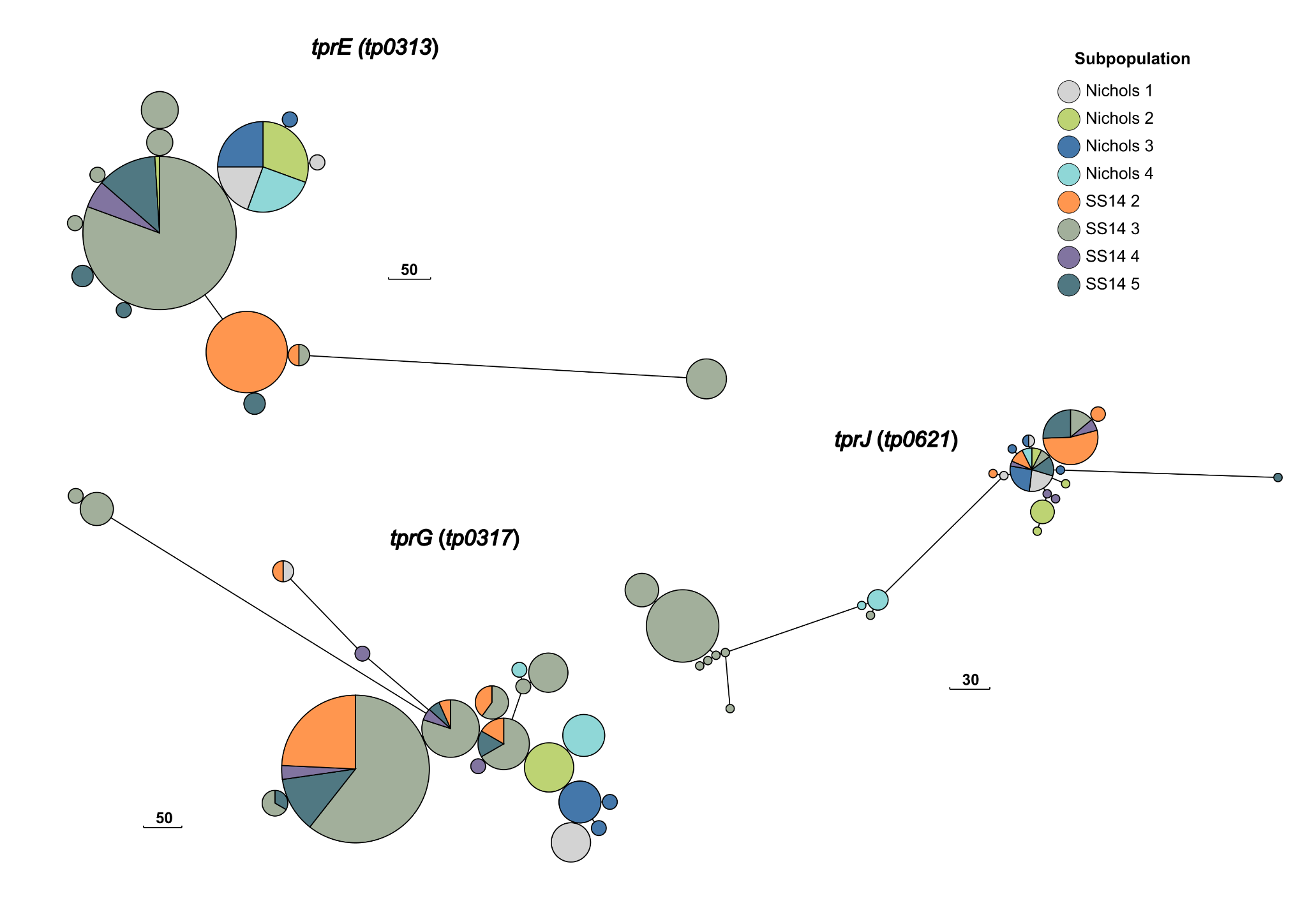

**Supplementary Figure 12**: A) Frequency of macrolide resistance-associated markers (A2058G/A2059G) in the TPA 23S rRNA gene among strains sequenced in this study. B) Frequency of the *tp0705* (*mrcA*) A1873G mutation, which has been associated with reduced in vitro susceptibility to ceftriaxone and penicillin G (Pospíšilová et al., 2025). *A* indicates the susceptible allele; *G* indicates the reduced‑susceptibility allele.

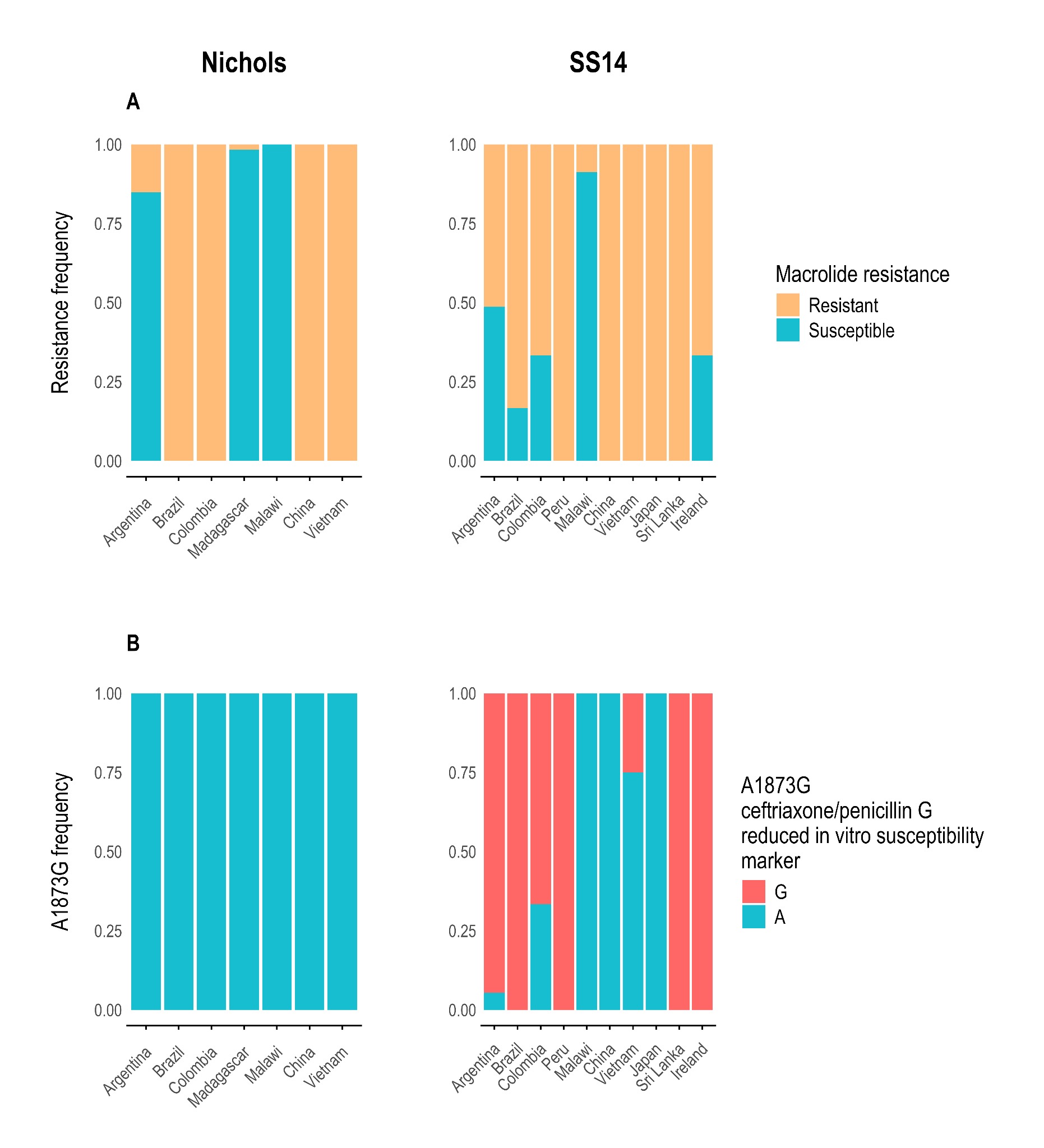
